# Bayesian Dose-Finding for Theta Burst Stimulation Tolerability: A Randomized Study Comparing Intermittent and Continuous Protocols at Distinct Prefrontal Targets

**DOI:** 10.64898/2026.08.10.26360147

**Authors:** George Kypriotakis, Lisa McTeague, Maher Karam-Hage, Brian A. Taylor, Sanjay Shete, Francesco Versace

## Abstract

**Background:** Theta burst stimulation (TBS) is an efficient form of repetitive transcranial magnetic stimulation, but tolerability may depend on target and stimulation pattern, limiting translation to accelerated protocols.

**Objective:** To estimate tolerable intensities for intermittent TBS (iTBS) over F3, approximating left dorsolateral prefrontal cortex, and continuous TBS (cTBS) over Fp1, intended to engage more anterior ventral/frontopolar circuitry, in non-treatment-seeking adults with obesity or tobacco use disorder.

**Methods:** In an open-label randomized crossover titration study, 64 adults completed two TBS visits 7 days apart. Each visit included 3 sessions of 600 pulses, beginning at 80% resting motor threshold (RMT) with protocol-permitted escalation or de-escalation. The primary endpoint was participant-level maximum final maintained intensity. Bayesian grouped-binomial logistic regression estimated the intensity tolerated by 70% of participants (ED_70_), and a prespecified rule selected the highest dose with at least 80% posterior probability of meeting 70% tolerability.

**Results:** Observed tolerability at 80% RMT was 86.4% for iTBS and 51.6% for cTBS. Primary-model ED_70_ was 105.5% RMT (95% credible interval [CrI], 99.0%-113.9%) for iTBS and 69.2% RMT (95% CrI, 65.0%- 73.4%) for cTBS. The recommended intensity was 100% RMT for iTBS and 60% RMT for cTBS; no cTBS dose at or above 80% RMT met criterion. cTBS produced greater immediate symptom burden, whereas 24-hour symptoms were uncommon.

**Conclusion:** iTBS over F3 supported a future-trial design window of 90%-100% RMT, whereas cTBS over Fp1 showed a tolerability ceiling below 80% RMT. Future cTBS protocols targeting ventral prefrontal circuitry may need to move dorsally to improve tolerability.

## Introduction

Repetitive transcranial magnetic stimulation (rTMS) is used for neuropsychiatric conditions including treatment-resistant depression and substance use disorders (SUDs) ^1^. By delivering repeated magnetic pulses through a scalp coil, rTMS induces cortical currents and modulates local and connected brain activity ^2^. Theta burst stimulation (TBS) delivers rTMS pulses in brief high-frequency bursts (50 Hz) repeated at theta frequency (5 Hz). Relative to conventional rTMS, TBS markedly reduces session duration: 3 minutes for intermittent TBS (iTBS) and about 40 seconds for continuous TBS (cTBS) ^3^. This favors accelerated protocols, which deliver multiple daily sessions and may produce rapid symptom improvement ^4,5^. However, accelerated protocols also magnify the practical importance of tolerability, because transient discomfort can become limiting with repeated same-day stimulation.

Many rTMS clinical interventions for psychiatric disorders target the dorsolateral prefrontal cortex (DLPFC)^6^. Left DLPFC stimulation is hypothesized to up-regulate executive control neurocircuitry ^7^. Accordingly, DLPFC stimulation has been proposed as a treatment for SUDs ^8^ and obesity ^9^. Accelerated iTBS to the DLPFC has shown preliminary promise for cocaine use disorder ^10^, smoking ^11^, and heroin use disorder ^12^. Stimulation of the ventromedial prefrontal cortex (VMPFC) has also been proposed to reduce drug-cue hyper-reactivity in SUDs ^13,14^. Pilot studies showed that cTBS to the VMPFC reduced cue-related brain responses and cravings in cocaine and alcohol use disorders ^13,15^.

It remains unclear whether TBS can be delivered with comparable tolerability at both cortical targets. iTBS to the left DLPFC is commonly delivered by positioning the coil at or near the F3 electrode site, whereas cTBS intended to engage ventral/frontopolar circuitry has used coil placement at Fp1, just above the left eyebrow^16^. Because skull-to-cortex distance is greater at Fp1 than at F3 ^17,18^, stimulation at Fp1 may require higher intensity to achieve comparable circuit engagement. However, higher intensity can reduce tolerability without improving clinical benefit: in a large depression trial targeting the left DLPFC, iTBS at 120% of resting motor threshold (RMT) caused greater stimulation-related discomfort than iTBS at 80% of RMT, with no difference in antidepressant efficacy ^19^. Although frontal pole stimulation is generally tolerated ^20^, the intensity range that can be reliably delivered at Fp1 versus F3 under specific TBS patterns remains unclear. Systematic tolerability data are essential for dose selection; future ventral/frontopolar trials could otherwise use intensities too low for target engagement or too high for consistent tolerability.

Bayesian dose-finding methods from oncology phase I trials, including the Continual Reassessment Method and Bayesian Optimal Interval design, provide a principled framework for this question ^21,22^. Unlike completion-rate summaries, these methods estimate dose-response relationships, quantify posterior uncertainty, and support prespecified probabilistic rules for future-trial dose selection. We applied this framework to a randomized crossover study of iTBS over F3 and cTBS over Fp1 in non-treatment-seeking adults with obesity or tobacco use disorder. F3 approximated left DLPFC^23^, and Fp1 was intended to engage left VMPFC^14^.

The goals were to (1) estimate the effective dose tolerated by at least 70% of participants (ED_70_) for each protocol, (2) apply a prespecified decision rule to recommend intensities for future trials, (3) compare protocols directly on tolerability at matched intensities, (4) characterize individual tolerability phenotypes, and (5) describe demographic correlates of tolerability and adverse event profiles.

## Methods

### Study Design and Participants

This randomized crossover titration study was analyzed using Bayesian dose-finding methods. Participants came from 2 prospective, non-treatment-seeking cohorts: the BRAIN study (n = 43), which enrolled adults with tobacco use disorder and the RAPID study (n = 21), which enrolled adults with obesity. Both studies were approved by the MD Anderson Cancer Center institutional review board (RAPID, 2024-0261, approved October 2, 2024, activated November 21, 2024; BRAIN, 2024-1316, approved January 30, 2025, activated February 19, 2025). All participants provided written informed consent. ClinicalTrials.gov identifiers were NCT06991062 (BRAIN) and NCT06639594 (RAPID). Eligible adults met standard transcranial magnetic stimulation safety criteria. Key exclusions were seizures or fainting spells, metallic implants above the neck, current pregnancy, and self-reported psychiatric disorders. The analytic dataset comprised 2010 titration observations from 64 unique participants (Table 1).

**Table 1.** Baseline Participant Characteristics.

| Characteristic | Overall (N = 64) |
| --- | --- |
| **Age, y** |  |
| Mean (SD) | 45.8 (8.8) |
| Range | 25–60 |
| **Sex at birth, No. (%)** |  |
| Male | 35 (54.7) |
| Female | 29 (45.3) |
| **Race, No. (%)** |  |
| Black or African American | 38 (59.4) |
| White | 21 (32.8) |
| Other | 5 (7.8) |
| **Ethnicity, No. (%)** |  |
| Hispanic or Latino | 10 (15.6) |
| Non-Hispanic | 54 (84.4) |
| **Study cohort, No. (%)** |  |
| BRAIN | 43 (67.2) |
| RAPID | 21 (32.8) |
| **Resting motor threshold** |  |
| Mean (SD) | 55.2 (12.1) |
| Range | 11–85 |

### Stimulation Protocols and Titration Endpoint

Two TBS protocols were delivered in a within-subject crossover design across 2 visits separated by a 7-day washout. At one visit, participants received intermittent TBS (iTBS) over F3; at the other, they received continuous TBS (cTBS) over Fp1. Visit order was randomized, with 33 participants assigned to iTBS first and 31 to cTBS first. Each visit comprised 3 sessions of 600 pulses separated by 15-minute intervals. Stimulation was delivered with a MagPro rTMS Research System and Cool-B65 figure-of-eight coil. Scalp positions corresponding to F3 and Fp1 were estimated using EEG-Locator software and registered in Brainsight neuronavigation software; resting motor threshold (RMT) was estimated using the TMS Motor Threshold Assessment Tool.

The nominal starting intensity was 80% RMT. Intensities could increase in 10% increments to 90%, 100%, and 110% RMT when tolerated, and could decrease below 80% RMT if participants reported intolerability. The primary tolerability endpoint was final maintained end-of-session intensity, not the highest transient intensity reached during titration. For each participant-protocol combination, the maximum session-final maintained intensity across the 3 sessions defined the individual maximum tolerated dose. This endpoint reflects the intensity future efficacy trials require participants to maintain.

### Statistical Analysis

The primary analysis used Bayesian grouped-binomial logistic regression to estimate protocol-specific tolerability across intensities. Dose was centered at 60% RMT and scaled in 10-percentage-point units, using the final maintained titration dose grid. The primary estimand was ED_70_, the intensity tolerated by at least 70% of participants. Dose selection followed a prespecified probabilistic rule: select the highest intensity for which the posterior probability of at least 70% tolerability was at least 80%. A clinical decision lookup table for alternative target and confidence combinations is provided in eTable 2.

Complementary analyses included mixed-effects logistic regression with participant random intercepts, grouped spline models, Bayesian isotonic regression, prior sensitivity analyses, and direct protocol-comparison models. Sequence, carryover, within-subject concordance, within-day session effects, and demographic subgroup analyses were secondary or exploratory and are detailed in the Supplementary Methods, eTables 1-8 and 11-14, and eFigures 1-4.

### Safety and Adverse Event Analysis

Participant-rated symptoms were assessed immediately after each TMS session and approximately 24 hours after each visit. Immediate assessments rated headache, pain, skin or scalp irritation, facial twitching, fatigue, and fearfulness or anxiety during stimulation and at the time of post-session assessment, using an 11-point scale from 0 to 10. Participants could also report up to 2 additional discomfort symptoms. The 24-hour TBS screener assessed delayed physical symptoms, mood/activation items, and additional symptoms not captured by structured items. A symptom was present if rating was greater than 0. Immediate moderate-or-greater symptoms were defined as ratings of 4 or higher, severe symptoms as ratings of 7 or higher, and 24-hour moderate-or-greater symptoms as ratings of 3 or higher on the 0 to 5 physical-symptom scale. Expanded scoring definitions are in the Supplementary Methods.

### Implementation

Analyses were conducted in R using brms for Bayesian regression and Stan for posterior sampling. Convergence was assessed using R-hat, effective sample size, divergent transitions, and posterior predictive checks; complete diagnostics are provided in the Supplementary Methods and eFigure 3.

## Results

### Participant Characteristics and Flow

Baseline characteristics of the 64 participants are shown in Table 1. Mean (SD) age was 45.8 (8.8) years (range, 25-60); 35 participants (54.7%) were male, 38 (59.4%) were Black or African American, 21 (32.8%) were White, and 10 (15.6%) identified as Hispanic or Latino. The BRAIN study contributed 43 participants (67.2%) and the RAPID study contributed 21 (32.8%). Full demographic details including expanded race categories are provided in eTable 9.

Of 64 participants, 59 contributed evaluable iTBS data, 62 contributed evaluable cTBS data, and 57 (89.1%) completed both protocols (eTable 10). Sequence allocation was balanced (33 iTBS-first, 31 cTBS-first). The mean number of sessions per participant-protocol was 2.98, and 99.2% of participant-protocol units completed all 3 planned sessions. The mean (SD) resting motor threshold was 55.2% (12.1%) of maximum stimulator output (range, 11%-85%).

### Observed Tolerability

Observed tolerability differed markedly between protocols across the full intensity range (Table 2; Figure 1). For iTBS targeting F3, tolerability remained high: 100% at 10-30% RMT, declining gradually to 98.3% at 40-50% RMT, 93.2% at 70% RMT, 86.4% at the protocol starting dose of 80% RMT, 78.0% at 90% RMT, 74.6% at 100% RMT, and 71.2% at the maximum tested intensity of 110% RMT. More than 70% of participants tolerated even the highest intensity tested.

**Figure 1.**
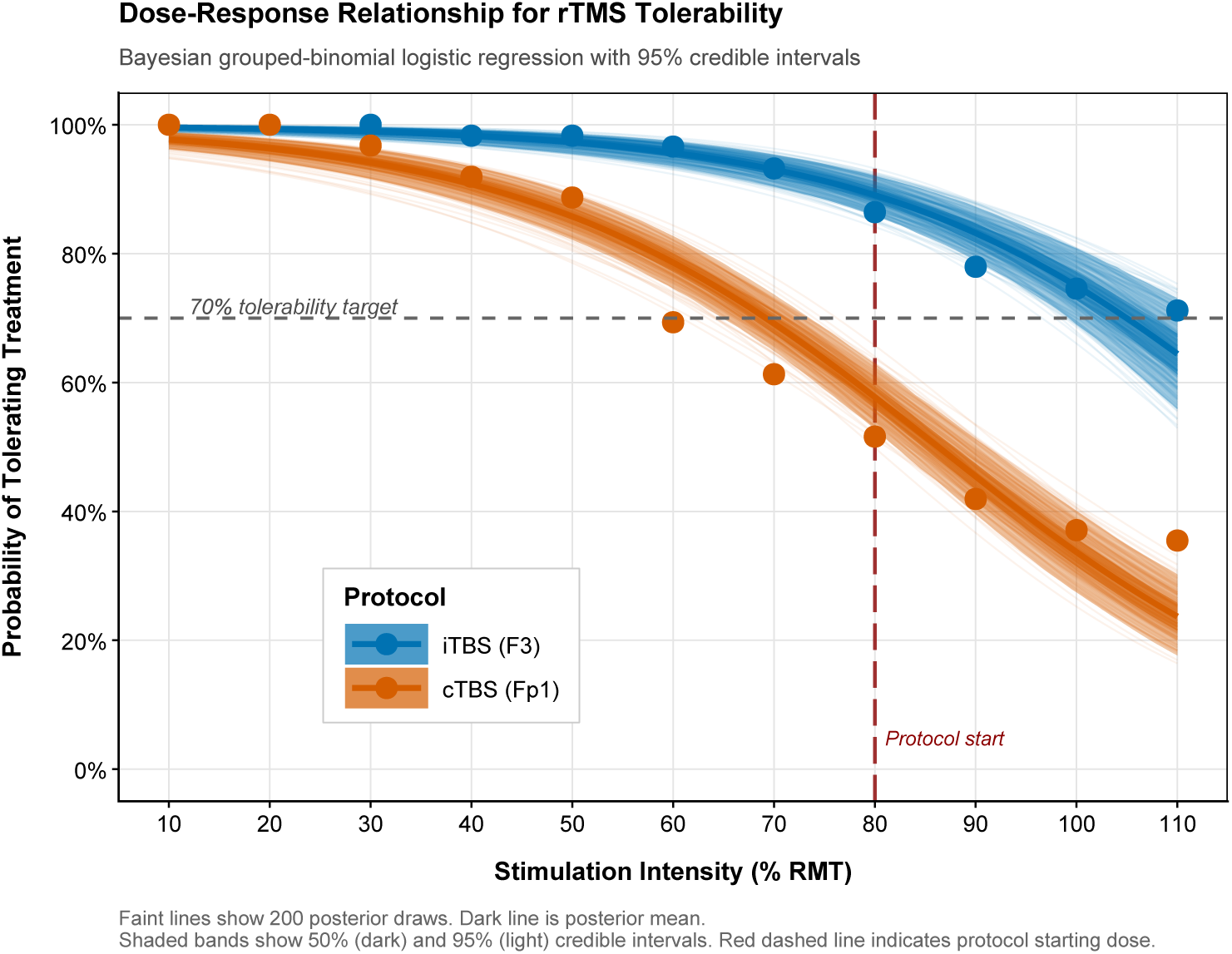
Bayesian grouped-binomial logistic regression estimates for iTBS at F3 (blue) and cTBS at Fp1 (orange). Points indicate observed proportions at each intensity threshold. Solid curves represent posterior mean estimates; dark and light bands indicate 50% and 95% credible intervals, respectively. Faint lines show 200 individual posterior draws illustrating parametric uncertainty. Horizontal dashed line marks the 70% tolerability target. Vertical red dashed line marks the protocol starting dose (80% RMT). The curves demonstrate a gradual, nearly linear decline for iTBS compared with a steeper logistic decline for cTBS, with the cTBS curve crossing the 70% target well below 80% RMT.

**Table 2.**
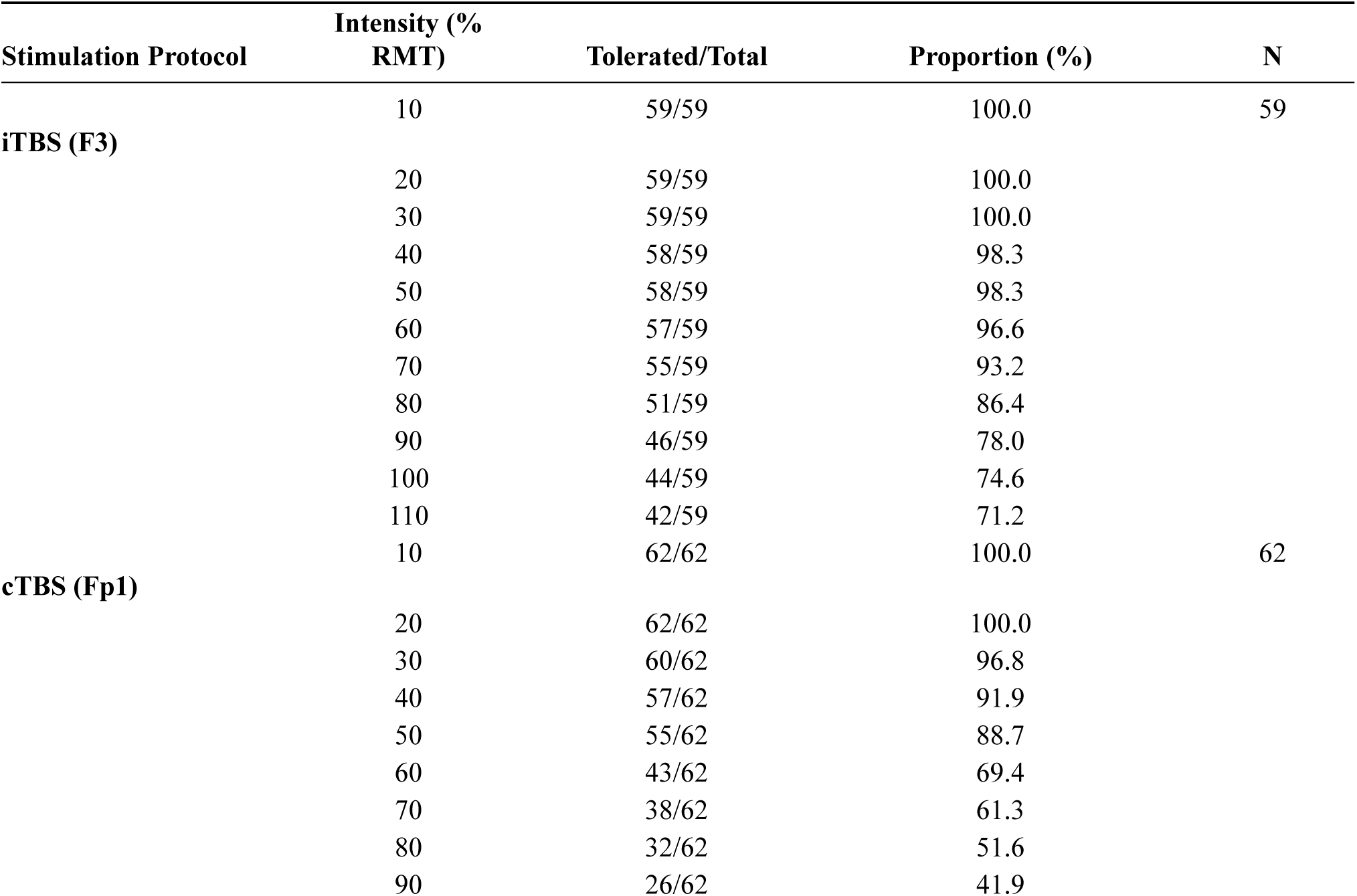
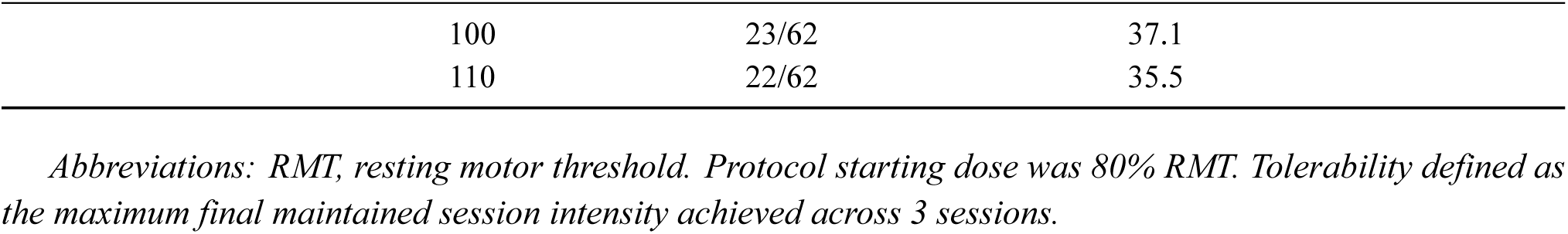
Observed Tolerability by Stimulation Intensity and Protocol.

| Stimulation Protocol | Intensity (% RMT) | Tolerated/Total | Proportion (%) | N |
| --- | --- | --- | --- | --- |
| iTBS (F3) | 10 | 59/59 | 100.0 | 59 |
|  | 20 | 59/59 | 100.0 |  |
|  | 30 | 59/59 | 100.0 |  |
|  | 40 | 58/59 | 98.3 |  |
|  | 50 | 58/59 | 98.3 |  |
|  | 60 | 57/59 | 96.6 |  |
|  | 70 | 55/59 | 93.2 |  |
|  | 80 | 51/59 | 86.4 |  |
|  | 90 | 46/59 | 78.0 |  |
|  | 100 | 44/59 | 74.6 |  |
|  | 110 | 42/59 | 71.2 |  |
| cTBS (Fp1) | 10 | 62/62 | 100.0 | 62 |
|  | 20 | 62/62 | 100.0 |  |
|  | 30 | 60/62 | 96.8 |  |
|  | 40 | 57/62 | 91.9 |  |
|  | 50 | 55/62 | 88.7 |  |
|  | 60 | 43/62 | 69.4 |  |
|  | 70 | 38/62 | 61.3 |  |
|  | 80 | 32/62 | 51.6 |  |
|  | 90 | 26/62 | 41.9 |  |
|  | 100 | 23/62 | 37.1 |  |
|  | 110 | 22/62 | 35.5 |  |

*Abbreviations: RMT, resting motor threshold. Protocol starting dose was 80% RMT. Tolerability defined as the maximum final maintained session intensity achieved across 3 sessions.*

For cTBS targeting Fp1 tolerability declined more steeply: 100% at 10-20% RMT, dropping to 91.9% at 40% RMT, 69.4% at 60% RMT, and only 51.6% at the protocol starting dose of 80% RMT. At 90% RMT, fewer than half (41.9%) tolerated cTBS, and at 110% RMT only 35.5% tolerated treatment. The divergence between protocols was already apparent by 50% RMT (98.3% vs 88.7%) and widened progressively at higher intensities.

### Primary Model Results and Dose Recommendations

The primary Bayesian grouped-binomial logistic regression demonstrated excellent convergence (all R-hat <1.01; minimum effective sample size, 2746; no divergent transitions; see eMethods). For iTBS, the ED_70_, the intensity tolerated by at least 70% of the patients, was 105.5% RMT (95% CrI, 99.0%-113.9%). Model-estimated tolerability was 89.1% at 80% RMT (95% CrI, 86.0%-91.9%), 83.3% at 90% RMT (95% CrI, 79.1%-87.0%), 75.1% at 100% RMT (95% CrI, 68.8%-80.7%), and 64.5% at 110% RMT (95% CrI, 55.3%-73.2%). Complete posterior estimates at every intensity level appear in eTable 11.

For cTBS, the ED_70_ was substantially lower at 69.2% RMT (95% CrI, 65.0%-73.4%), falling below the protocol starting dose of 80% RMT. Model-estimated tolerability at 80% RMT was only 57.8% (95% CrI, 52.8%- 62.7%), declining to 45.5% at 90% RMT and 33.8% at 100% RMT. The posterior distributions of ED_70_ for the two protocols were well separated, consistent with markedly different tolerability profiles. At each protocol-relevant dose, the number of patients expected to tolerate treatment per 100 treated is provided in eTable 12.

Applying the prespecified decision rule, the recommended intensity was 100% RMT for iTBS, where the posterior probability of achieving at least 70% tolerability was 94.5%. At 110% RMT, this probability fell to 12.0%, narrowly missing the 80% decision threshold. For cTBS, the recommended intensity was 60% RMT (posterior probability, 99.98%). No cTBS intensity at or above 80% RMT met the decision criterion; at 70% RMT, the probability was only 35.0%.

### Sensitivity and Robustness Analyses

All complementary and robustness analyses preserved the finding that iTBS has a wider tolerability window than cTBS (Table 3; Figure 2). In the mixed-effects model, ED_70_ estimates were 112.8% RMT (95% CrI, 100.8%- 127.1%) for iTBS and 76.2% RMT (95% CrI, 66.9%-85.1%) for cTBS. The wider credible intervals reflected the additional variance absorbed by participant random intercepts, and marginal recommendations were more conservative: 90% RMT for iTBS and 50% RMT for cTBS. Grouped spline models recommended 100% RMT for iTBS and 60% RMT for cTBS, while Bayesian isotonic regression supported escalation through 110% RMT for iTBS but no acceptable cTBS dose. Thus, the grouped and mixed models support a practical iTBS design window of 90%-100% RMT, while no cTBS dose at or above 80% RMT met criterion. Prior sensitivity was stable (eTable 1). The simulation study showed comparable or superior correct selection rates relative to the 3+3 design, with advantages in low-tolerability scenarios resembling cTBS (eMethods).

**Figure 2.**
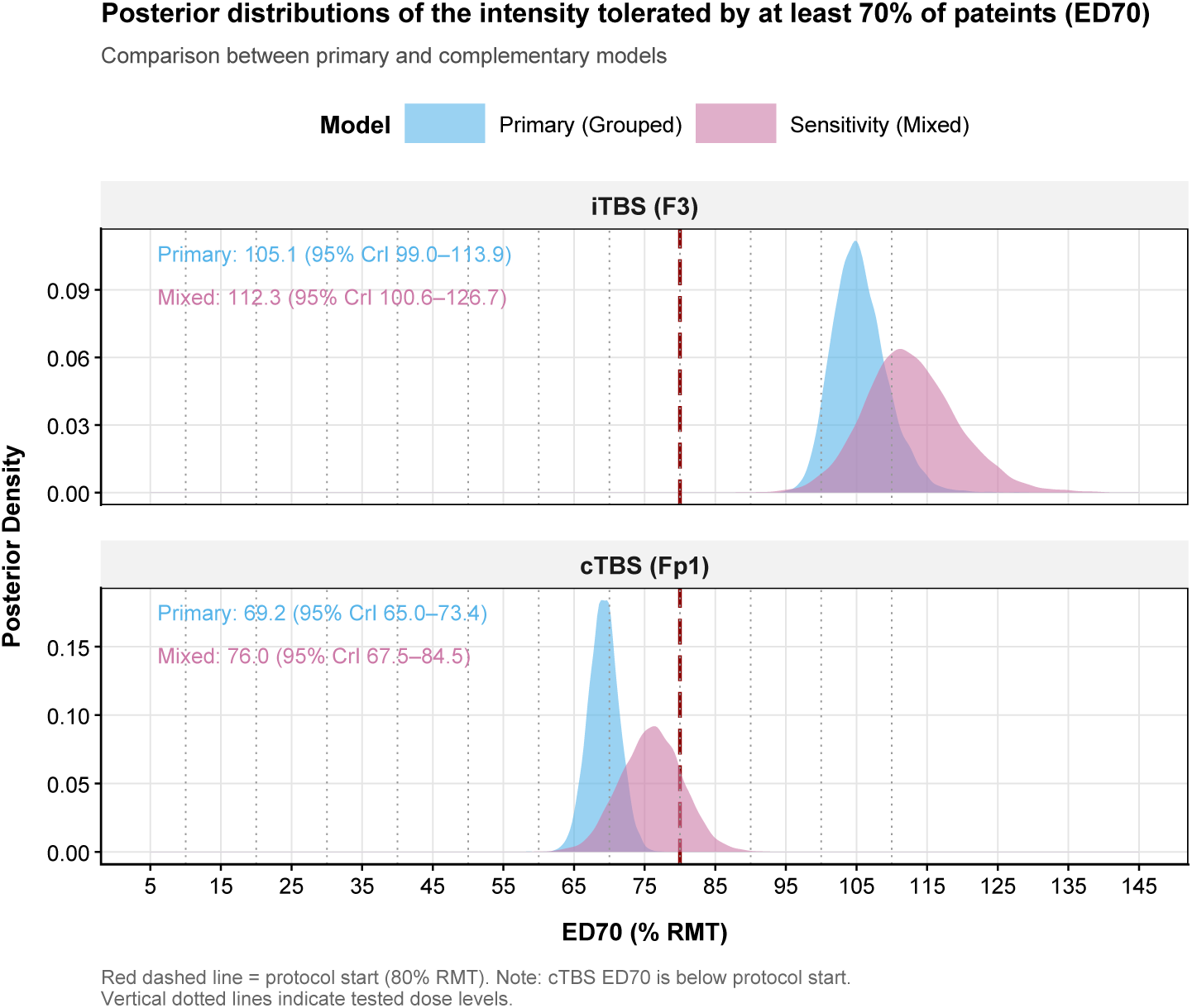
ED_70_ Estimates Across Statistical Models. Distributions comparing ED_70_ posterior means and 95% credible intervals from the primary grouped-binomial and mixed-effects complementary models for each protocol. Blue symbols represent iTBS; orange symbols represent cTBS. The red dashed line marks the protocol starting dose (80% RMT). The cTBS ED_70_ falls below 80% RMT across both models, confirming that no standard therapeutic intensity meets the 70% tolerability target.

**Table 3.** Dose-Finding Results Across Statistical Models.

| Protocol | Model | $ED_{70}$ (95% CrI) | $P(\geq 70\% \text{ at Rec. Dose})$ | Recommended Intensity |
| --- | --- | --- | --- | --- |
| **iTBS (F3)** | Grouped logistic (primary) | 105.5 (99.0–113.9) | 94.5% | 100% RMT |
|  | Mixed-effects (complementary) | 112.8 (100.8–127.1) | 85.5% | 90% RMT |
|  | Spline (robustness) | - | 94.3% | 100% RMT |
|  | Isotonic (robustness) | - | 88.4% | 110% RMT |
| **cTBS (Fp1)** | Grouped logistic (primary) | 69.2 (65.0–73.4) | 99.9% | 60% RMT |
|  | Mixed-effects (sensitivity) | 76.2 (66.9–85.1) | 92.7% | 50% RMT |
|  | Spline (robustness) | - | 91.5% | 60% RMT |
|  | Isotonic (robustness) | - | - | None |

*Abbreviations: $ED_{70}$ , intensity tolerated by at least 70% of the patients; CrI, credible interval; RMT, resting motor threshold. Protocol start = 80% RMT. Spline and isotonic models do not yield closed-form $ED_{70}$ .*
*Note: $P(\geq 70\% \text{ at Rec. Dose})$ is the posterior probability of achieving the tolerability target at the model-specific recommended dose. In the mixed-effects model, $ED_{70}$ is conditional on the random effect being 0, whereas recommendation probabilities are marginalized over the random-effect distribution*

### Direct Protocol Comparison

The joint protocol-comparison model (eTable 13 for full results) showed consistently higher tolerability for iTBS than cTBS at every tested intensity. The protocol main effect was −1.82 on the logit scale (95% CrI, −2.37 to −1.30), indicating substantially lower baseline tolerability for cTBS. The dose-by-protocol interaction was negligible (0.009; 95% CrI, −0.14 to 0.17), indicating that the slopes of decline were similar and the iTBS advantage was driven primarily by a higher intercept.

At the protocol starting dose of 80% RMT, estimated tolerability was 89.2% for iTBS versus 58.0% for cTBS, a difference of 31.2 percentage points (95% CrI, 25.6-36.8 pp; posterior probability P[iTBS > cTBS] = 100%). At 100% RMT, the difference widened to 41.2 percentage points (95% CrI, 32.4-49.6 pp). Even at 50% RMT, iTBS showed an 11.5 percentage-point advantage (95% CrI, 7.7-15.3 pp). At every intensity from 30% to 110% RMT, the posterior probability that iTBS tolerability exceeded cTBS was at least 99.9%.

### Sequence Effects and Within-Subject Concordance

Analysis of treatment sequence effects revealed no clinically meaningful differences for cTBS (eTable 4; eFigure 4). For iTBS, tolerability estimates were similar regardless of sequence at low to moderate intensities, with some divergence at very high intensities (≥100% RMT) that did not alter dose recommendations. Carryover effects were not statistically significant (eTable 5): neither prior iTBS exposure (estimate, -3.87% RMT; 95% CrI, -17.84 to 9.79) nor prior cTBS exposure (6.18% RMT; 95% CrI, -7.66 to 19.93) meaningfully influenced second-period tolerability. Session-level changes were modest within the single-day accelerated protocol. For cTBS, session-level analyses did not suggest habituation; instead, the estimated session effect was negative, consistent with a modest within-day decline in maximum tolerated intensity across sessions (eTable 6; eFigure 1).

Among the 57 participants completing both protocols, within-subject concordance between iTBS and cTBS maximum tolerated intensities was only moderate (Pearson r, 0.590; 95% CI, 0.389-0.737) and absolute agreement was poor (ICC, 0.381; 95% CI, -0.028 to 0.650; eTable 3). Lin’s concordance correlation coefficient was 0.377 (95% CI, 0.221-0.513), confirming poor interchangeability. Bland-Altman analysis showed a mean difference of 22.6% RMT (iTBS higher) with wide limits of agreement (-23.7% to 69.0% RMT), indicating substantial individual variation in protocol-specific tolerability.

### Tolerability Phenotypes

Exploratory tolerability phenotypes are shown in Figure 3 and eTable 14. For iTBS, 44 participants (74.6%) were high tolerators (≥100% RMT), 7 (11.9%) were moderate tolerators (80%-99% RMT), and 8 (13.6%) were low tolerators (<80% RMT). For cTBS, 23 (37.1%) were high tolerators, 9 (14.5%) moderate tolerators, and 30 (48.4%) low tolerators. The cTBS low-tolerator group had a lower mean tolerated intensity than the iTBS low-tolerator group (51.0% vs 60.0% RMT) and a wider tolerated range (20%-70% vs 30%-70% RMT).

**Figure 3.**
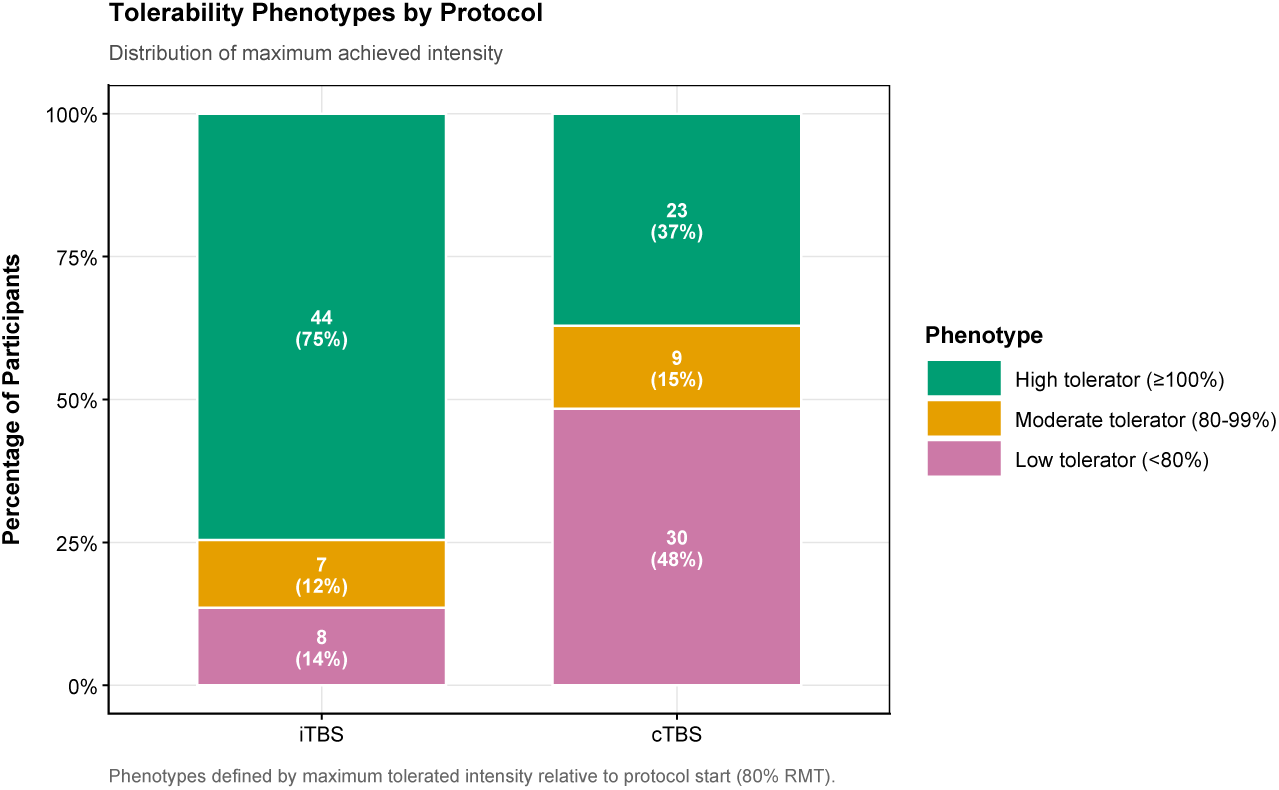
Distribution of Tolerability Phenotypes by Protocol. Participant-level tolerability phenotypes based on the maximum final maintained intensity achieved across sessions. High tolerators (green) reached at least 100% RMT; moderate tolerators (gold) reached 80% to 99% RMT; low tolerators (pink) did not reach 80% RMT. Numbers within bars indicate count and percentage. For iTBS (n = 59), three-quarters were high tolerators with only 13.6% classified as low tolerators. For cTBS (n = 62), nearly half were low tolerators and only 37.1% were high tolerators.

**Figure 4.**
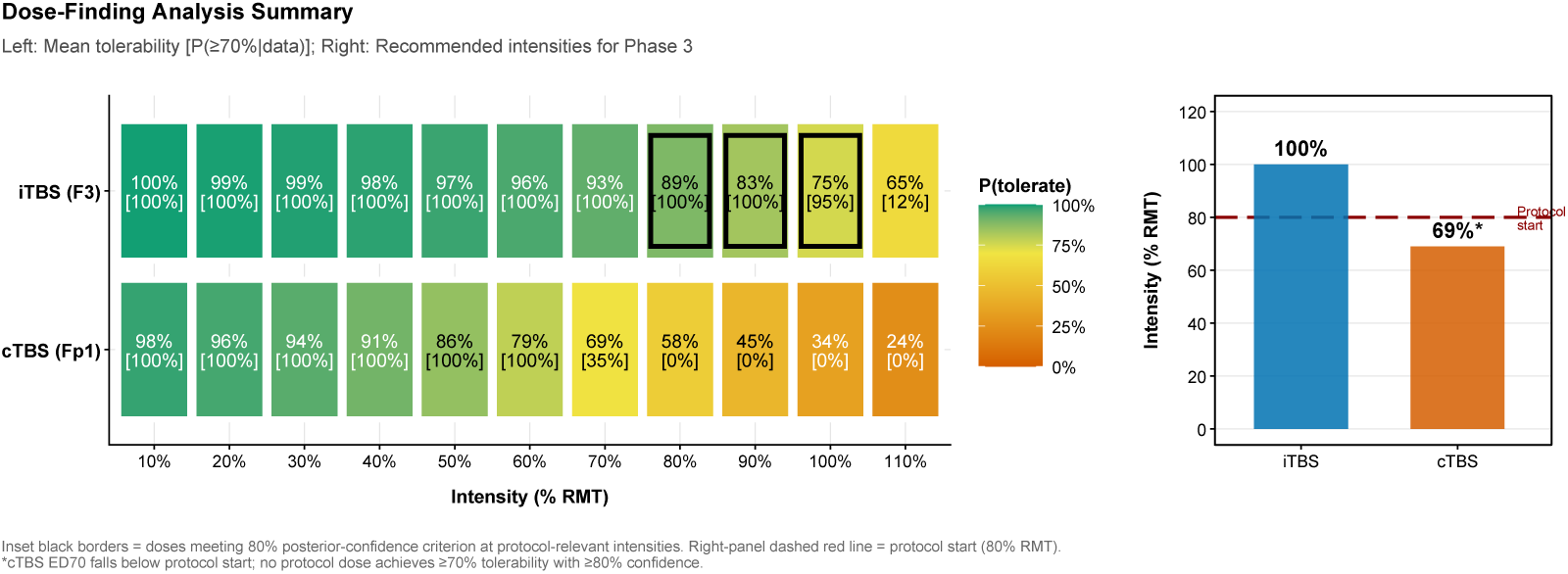
Integrated Dose-Finding Summary Dashboard. Left panel shows modeled tolerability (color scale) and the posterior probability of meeting the 70% target (bracketed values) at each intensity for both protocols. Black borders highlight intensities meeting the decision criterion at protocol-relevant doses (≥80% RMT). Right panel shows the primary recommended starting intensities relative to the 80% RMT starting dose. iTBS is recommended at 100% RMT; the cTBS recommendation (60% RMT) falls below protocol start.

### Demographic Subgroup Analyses

Exploratory analyses of tolerability by sex, age group, race, and ethnicity are detailed in eTables 7 and 8 and eFigure 2. For iTBS, mean (SD) maximum tolerated intensity was similar between men (101.5% [16.8%] RMT) and women (97.3% [21.3%] RMT; Wilcoxon P = .395). For cTBS, however, men achieved a substantially higher mean maximum tolerated intensity (88.2% [27.8%] RMT) than women (64.3% [25.2%] RMT; P = .002), with 67.9% of women classified as low tolerators compared with 32.4% of men.

Tolerability did not differ significantly by age group or ethnicity for either protocol. Heterogeneity in iTBS tolerability across race categories was statistically significant (P = .039) but involved small cells and was exploratory; the race effect was not significant for cTBS (P = .130).

### Safety and Adverse Events

Overall, the symptom dataset included 125 participant-visit records from 65 participants. Immediate post-session symptom assessments were complete for 120 visits, all of which linked to protocol, yielding 360 analyzed sessions (177 iTBS sessions and 183 cTBS sessions). Rated 24-hour TBS screener assessments were available for 120 protocol-linked visits.

During rTMS, symptom occurrence differed by stimulation protocol. At the session level, the most common iTBS symptom was facial twitching (118/177, 66.7%). For cTBS, pain was the most common symptom (140/183, 76.5%), followed by facial twitching and skin or scalp irritation (Table 4; Figure 5; eTable 15). At the visit level, cTBS had a higher immediate symptom burden than iTBS: mean visit-level total burden was 52.6 for cTBS versus 32.1 for iTBS, and any moderate-or-greater immediate symptom occurred in 90.2% versus 69.5% of visits (eTable 16).

**Figure 5.**
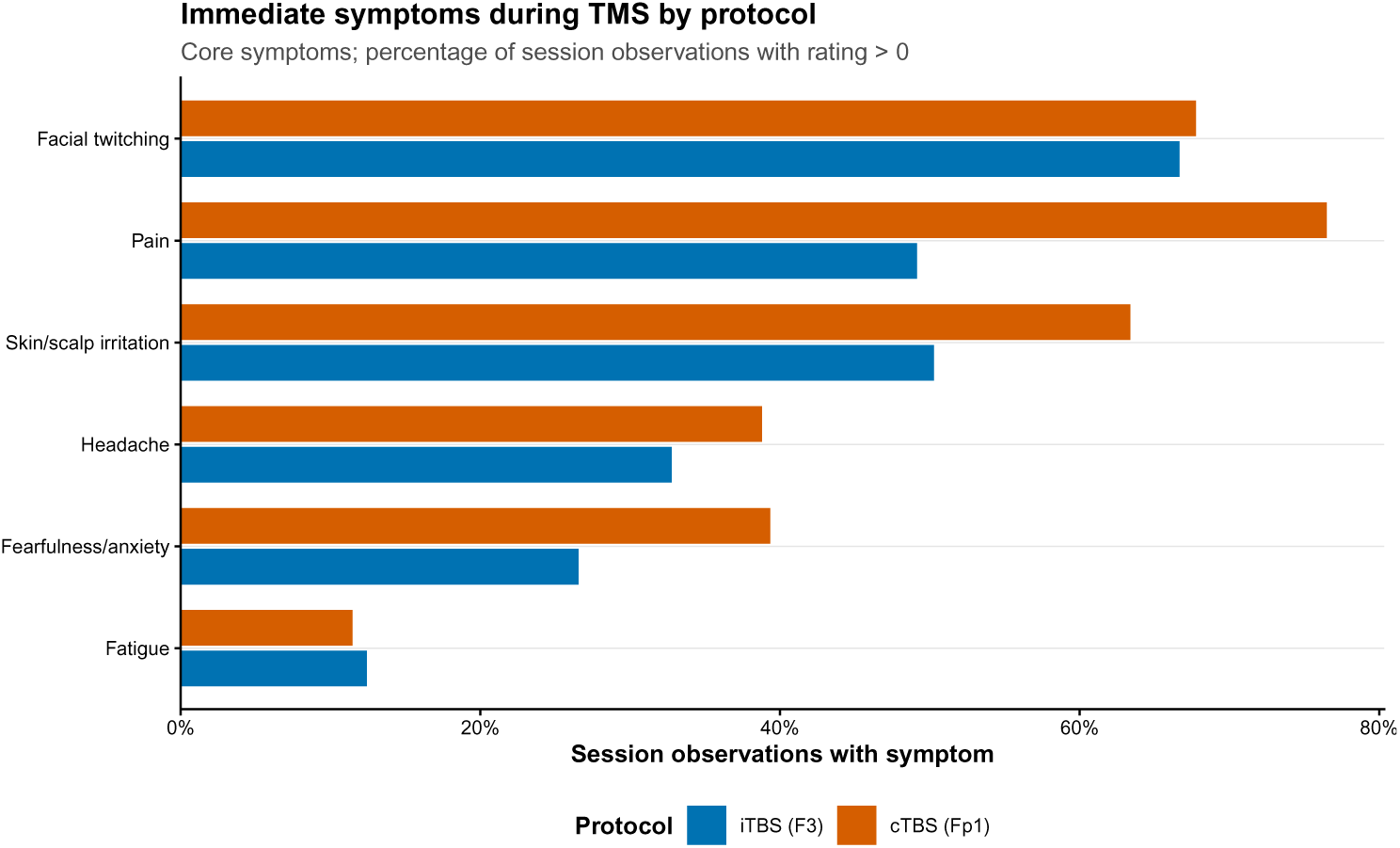
Participant-rated immediate symptom prevalence during TMS by protocol. Bars show the percentage of session-level observations with symptom rating greater than 0 for each core immediate symptom. Symptoms were rated immediately after each TMS session for symptoms experienced during stimulation. Denominators were 177 iTBS session observations and 183 cTBS session observations. Blue indicates iTBS (F3); orange indicates cTBS (Fp1).

**Table 4.**
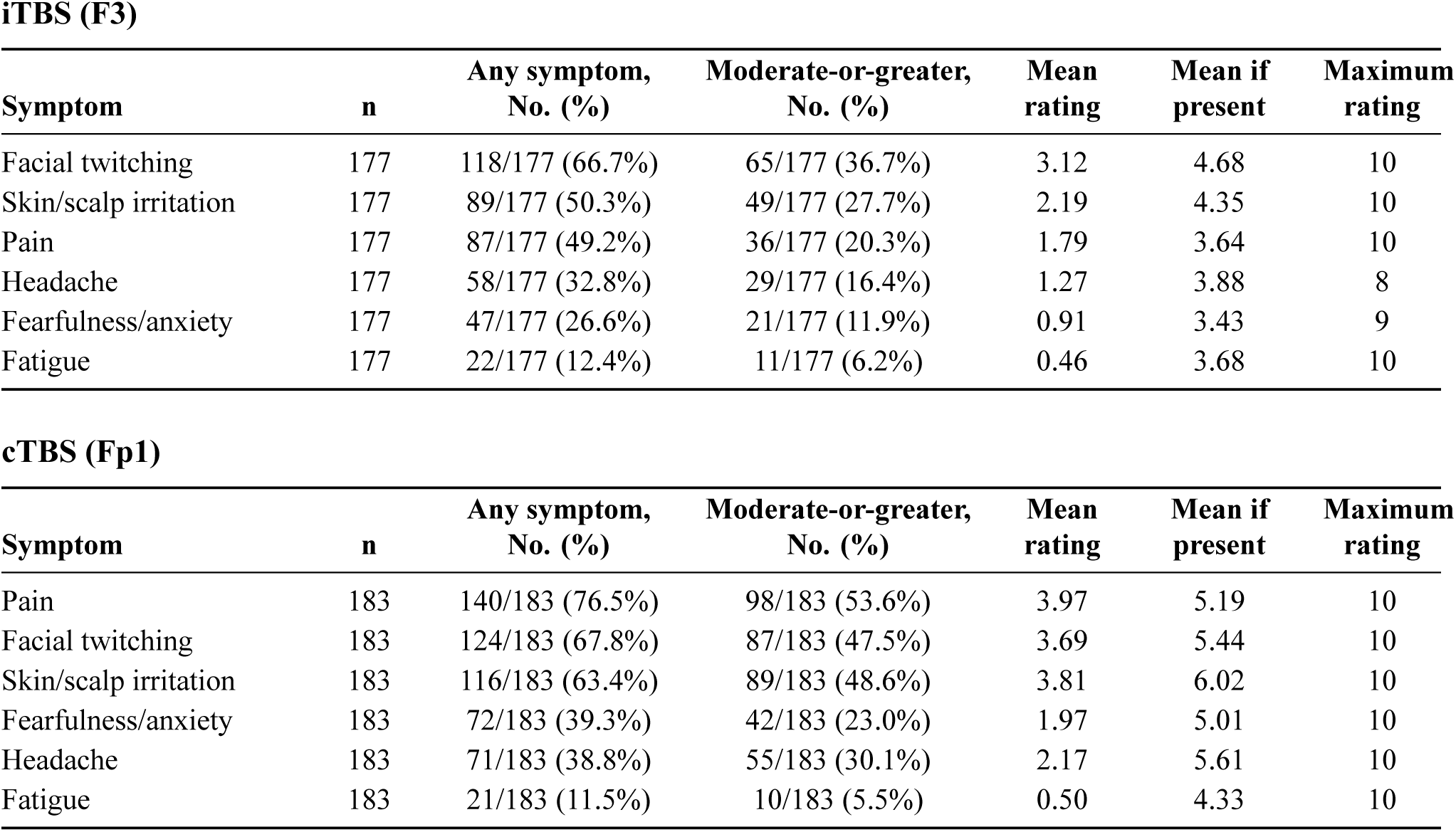
Participant-Rated Immediate Symptoms During TMS by Stimulation Protocol. *Moderate-or-greater is rating ≥4 on the 0 to 10 scale; n indicates analyzed sessions*.

Among 57 participants with data from both stimulation protocols, cTBS produced greater immediate symptom burden than iTBS. cTBS exceeded iTBS by a mean of 19.2 points in visit-level total burden (P < .001), 1.3 points in maximum single-session symptom severity rating (P < .001), and 2.7 moderate-or-greater symptom ratings per visit (P < .001; eTable 17). In a mixed model adjusted for final session intensity, session number, and visit, cTBS was associated with a 4.8-point higher immediate burden than iTBS (P < .001; eTable 18).

Immediate symptom burden declined across sessions within visits (Figure 6; eTable 19). Mean total burden decreased from session 1 to session 3 for iTBS (12.9 to 9.2) and cTBS (19.1 to 15.8). In adjusted mixed models, session 3 was associated with a 3.4-point lower total burden than session 1 (P < .001); the odds of any moderate-or-greater symptom were also lower at session 3 (OR, 0.19; P < .001). Visit 2 had lower total burden than visit 1 (estimate, -2.1; P = .012).

**Figure 6.**
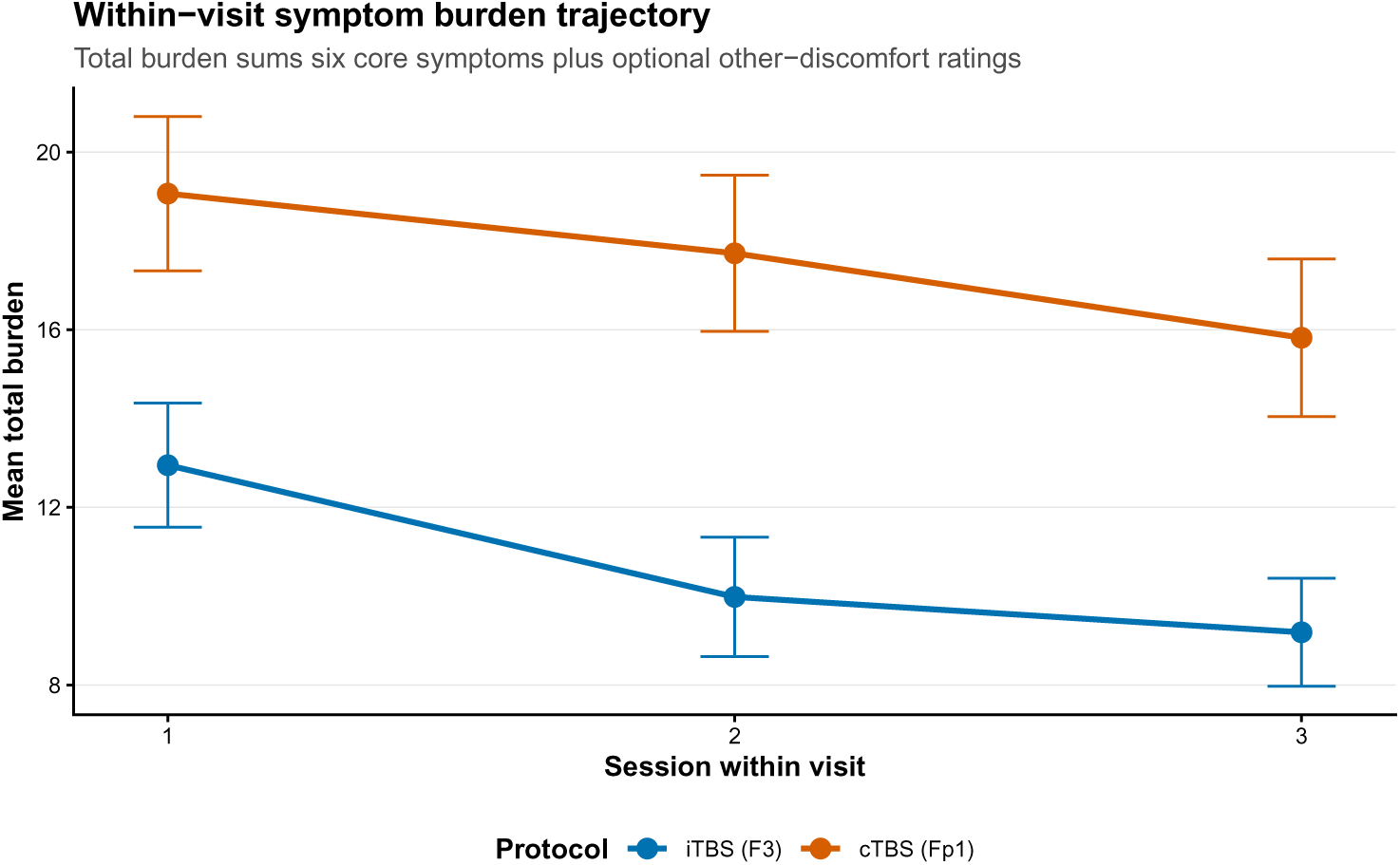
Mean total immediate symptom burden across the 3 sessions within each visit. Points show mean session-level total symptom burden across sessions 1, 2, and 3 within each visit; error bars indicate standard errors. Session-level total symptom burden was calculated as the sum of the 6 core during-TMS symptom ratings plus any rated other-discomfort items within that session. Higher values indicate greater symptom number, severity, or both. Blue indicates iTBS (F3); orange indicates cTBS (Fp1).

Most symptoms present during TMS improved immediately after the session, but resolution varied by symptom (eTable 20; eFigure 5). Facial twitching resolved post-session in 94.9% of iTBS occurrences and 85.5% of cTBS occurrences. Pain resolved in 71.3% and 70.7%, respectively. Headache and fatigue were more likely to persist at the immediate post-session assessment than the other core symptoms.

At the 24-hour follow-up, symptom prevalence was lower than during-session symptom prevalence (Table 5; Figure 7; eTable 21). Headache was the most common 24-hour physical symptom (12/59, 20.3% after iTBS; 11/61, 18.0% after cTBS). Other 24-hour symptoms were infrequent, although weakness was reported more often after cTBS than iTBS. Mood/activation items were also uncommon; unusual energy or motivation was reported in 11.9% of iTBS visits and 9.8% of cTBS visits (eTable 22).

**Figure 7.**
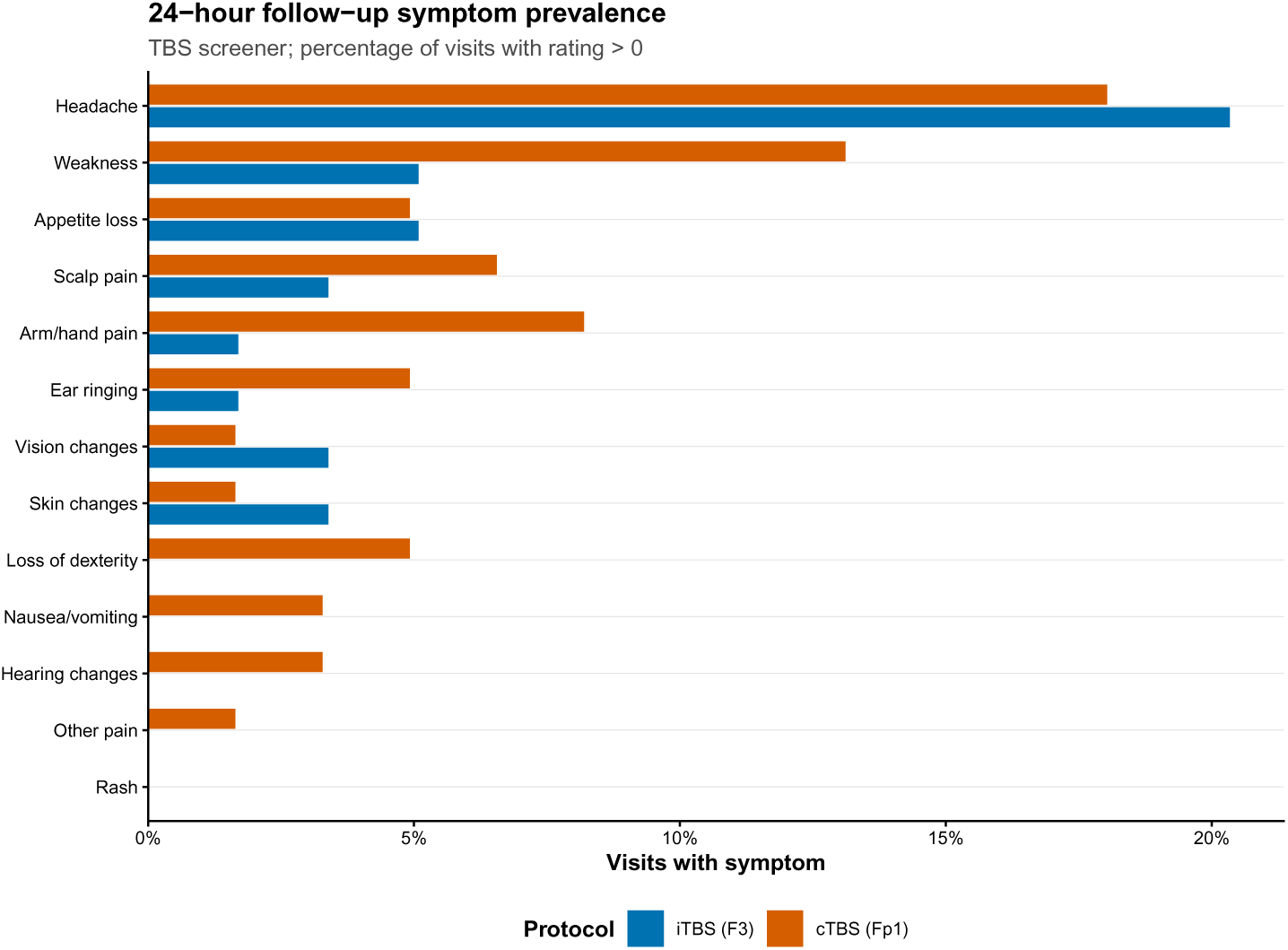
24-hour TBS screener physical symptom prevalence by protocol. Bars show the percentage of protocol-linked visits with a 24-hour TBS screener physical symptom rating greater than 0. Denominators were 59 iTBS visits and 61 cTBS visits with rated 24-hour physical symptom data. Only symptoms with nonzero prevalence in at least 1 protocol are displayed. Blue indicates iTBS (F3); orange indicates cTBS (Fp1).

**Table 5.** 24-Hour TBS Screener Physical Symptoms by Stimulation Protocol.

iTBS (F3)
| Symptom | Visits, No. | Any symptom, No. (%) | Mean rating | Maximum rating |
| --- | --- | --- | --- | --- |
| Headache | 59 | 12/59 (20.3%) | 0.44 | 5 |
| Appetite loss | 59 | 3/59 (5.1%) | 0.20 | 5 |
| Weakness | 59 | 3/59 (5.1%) | 0.19 | 5 |
| Scalp pain | 59 | 2/59 (3.4%) | 0.03 | 1 |
| Skin changes | 59 | 2/59 (3.4%) | 0.05 | 2 |
| Vision changes | 59 | 2/59 (3.4%) | 0.14 | 5 |
| Arm/hand pain | 59 | 1/59 (1.7%) | 0.02 | 1 |
| Ear ringing | 59 | 1/59 (1.7%) | 0.05 | 3 |

cTBS (Fp1)
| Symptom | Visits,<br>No. | Any symptom,<br>No. (%) | Mean<br>rating | Maximum<br>rating |
| --- | --- | --- | --- | --- |
| Headache | 61 | 11/61 (18.0%) | 0.34 | 5 |
| Weakness | 61 | 8/61 (13.1%) | 0.26 | 5 |
| Arm/hand pain | 61 | 5/61 (8.2%) | 0.21 | 4 |
| Scalp pain | 61 | 4/61 (6.6%) | 0.18 | 5 |
| Appetite loss | 61 | 3/61 (4.9%) | 0.18 | 5 |
| Ear ringing | 61 | 3/61 (4.9%) | 0.07 | 2 |
| Loss of dexterity | 61 | 3/61 (4.9%) | 0.11 | 4 |
| Hearing changes | 61 | 2/61 (3.3%) | 0.11 | 4 |
| Nausea/vomiting | 61 | 2/61 (3.3%) | 0.08 | 4 |
| Other pain | 61 | 1/61 (1.6%) | 0.02 | 1 |
| Skin changes | 61 | 1/61 (1.6%) | 0.02 | 1 |
| Vision changes | 61 | 1/61 (1.6%) | 0.08 | 5 |

*Table 5 summarizes the rated 24-hour physical symptoms; the 3 yes/no mood/activation items from the TBS screener are summarized separately in eTable 22 because they used a different response format. Only symptoms with nonzero prevalence in at least 1 protocol are shown*.

Optional other-discomfort reports were entered during 23.2% of iTBS sessions and 28.4% of cTBS sessions (eTables 23 and 24). Descriptions most often involved teeth or jaw sensations, eye watering or eye vibration, tapping or shock-like scalp sensations, sound irritation, nasal or sinus discomfort, vision changes, tinnitus, and arm or hand sensations.

## Discussion

In standardized titration data from 64 non-treatment-seeking adults with obesity or tobacco use disorder undergoing accelerated TBS, we identified a substantial and robust tolerability difference between iTBS to F3 and cTBS to Fp1. Under the prespecified rule, iTBS supported escalation to 100% RMT, while cTBS did not support any dose at or above the 80% RMT starting intensity. These findings, consistent across grouped-binomial, mixed-effects, spline, and isotonic models, inform future TBS trial design, particularly accelerated protocols targeting prefrontal circuits implicated in addictive and maladaptive appetitive behavior^24^

The primary finding, that iTBS ED_70_, the intensity tolerated by 70% of participants, exceeds 100% RMT while cTBS ED_70_ falls near 69% RMT, represents an approximately 37% RMT difference in tolerated intensity between the two protocol-target combinations. Clinically, iTBS can be delivered at F3 within the therapeutic range used in depression, while cTBS at Fp1 faces a ceiling below conventional therapeutic intensity. The joint model confirmed a 31 percentage-point tolerability advantage for iTBS at 80% RMT, growing to 41 percentage points at 100% RMT, with posterior certainty exceeding 99.9% at every tested intensity.

The complementary mixed-effects model recommended a more conservative iTBS intensity (90% vs 100% RMT), reflecting between-subject heterogeneity in population-averaged predictions. Together, the models suggest a practical iTBS design window of 90%-100% RMT, depending on tolerance for uncertainty, attrition, and implementation constraints. This range is consistent with evidence that sub-threshold iTBS (80% RMT) can produce clinical effects similar to supra-threshold iTBS (120% RMT) in depression despite less discomfort ^19^, suggesting that tolerability may determine the useful intensity ceiling. The clinical decision lookup table (eTable 2) supports alternative target and confidence choices.

The cTBS findings warrant caution because cTBS at Fp1 has been proposed to reduce cue reactivity and craving in SUDs ^14,15,25^. Across analyses, however, cTBS at Fp1 failed to meet the 70% tolerability target at or above 80% RMT. This constraint may reflect stimulation near the frontal sinuses, orbital structures, and supraorbital nerves, potentially amplified by the continuous burst pattern. Lowering intensity may improve comfort but could compromise target engagement given greater Fp1 scalp-to-cortex distance ^17,18^.

Thus, cTBS protocols intended to engage ventral/frontopolar circuitry may require protocol modifications, alternative coil positioning, or recalibrated tolerability targets before efficacy testing. Alternative coil positioning is plausible because lesion-network mapping identified addiction-relevant hubs in medial fronto-polar cortex and paracingulate gyrus ^26^, supporting future testing of more dorsal midline frontal targets, such as Fz/FCz or neuronavigation-guided medial prefrontal placements, that may preserve circuit relevance while improving tolerability.

Tolerability phenotypes revealed substantial individual variation. Three-quarters of iTBS recipients achieved ≥100% RMT, but 13.6% could not sustain 80% RMT; for cTBS, nearly half were low tolerators. This pattern is consistent with inter-individual variability in TBS neurophysiological effects ^27,28^ and suggests tolerability phenotyping may be as important as dose selection. Poor within-subject concordance also means high iTBS tolerability cannot be assumed to predict cTBS tolerability at Fp1.

Using final maintained intensity rather than transient intra-session peak targeted what can be delivered reliably within a session. This matters for Phase 3 planning, where participants must sustain target intensity to receive the intended dose. In the current data, 56% of cTBS sessions ended below their intra-session maximum, versus 27% for iTBS, confirming that transient tolerance is a poor proxy for sustained tolerability, particularly for cTBS.

Exploratory demographic findings suggest cTBS tolerability may be lower among women than men (P = .002), with two-thirds of women classified as low tolerators for cTBS compared with one-third of men. This finding warrants replication; potential mechanisms include scalp-to-cortex distance, pain perception, and hormonal influences on neural excitability. The race heterogeneity for iTBS (P = .039) should be interpreted cautiously given small cells and lack of conservative multiplicity correction. Tolerability did not differ by age group or ethnicity for either protocol, supporting broad applicability of the dose recommendations.

Together, symptom and dose-finding data suggest protocol tolerability was limited mainly by acute discomfort, not persistent adverse effects. cTBS delivered at Fp1 produced greater immediate symptom burden, especially pain, skin/scalp irritation, facial twitching, and fearfulness/anxiety. Most symptoms resolved by immediate post-session assessment, and 24-hour symptoms were uncommon; headache rates were similar after iTBS and cTBS. Thus, iTBS tolerability appears to reflect a wider acute comfort and dose-escalation margin, not lower delayed symptom burden. This matters for accelerated or multi-session protocols, where repeated same-day sessions can make acute discomfort dose-limiting.

Bayesian dose-finding improves on completion-rate summaries by estimating dose-response, quantifying posterior uncertainty, and applying transparent decision rules. Adapted from model-based oncology approaches, it formalizes risk-benefit tradeoffs and reproducibly compares candidate stimulation intensities in other neuromodulation tolerability questions ^21,22^. For early-phase neuromodulation, it translates titration data into probability-based intensity recommendations rather than observed completion rates or adverse-event counts alone.

Limitations include participant-reported tolerability during titration, limited demographic-subgroup precision, and lack of long-term tolerability data. The crossover design may have introduced sequence effects, although carryover analysis found none (eTable 5). Findings apply to these accelerated schedules, TBS patterns, and cortical targets and may not generalize to conventional rTMS protocols or alternative targets. Because participants were not seeking TBS treatment, absolute tolerability thresholds may differ in treatment-seeking populations willing to tolerate greater discomfort for expected clinical benefit. However, the relative tolerability disadvantage of cTBS at Fp1 likely remains relevant for treatment development, particularly accelerated protocols requiring repeated same-day stimulation. Although higher intensity is often presumed to enhance neuromodulatory effects, clinical dose-response relationships for these TBS protocols remain incompletely defined.

### Conclusions

In this Bayesian dose-finding analysis, iTBS to F3 showed a wide tolerability margin supporting escalation to 100% RMT under the prespecified rule, with a favorable safety profile across demographic subgroups. Mixed-effects modeling supported 90% RMT as a conservative iTBS option, suggesting a 90%-100% RMT design window for future trials. cTBS to Fp1, in contrast, had a persistent tolerability constraint, with no dose at or above the standard 80% RMT starting intensity meeting the prespecified criterion in any of the 4 analytical models. These findings support F3 for accelerated iTBS protocols and suggest cTBS protocols targeting ventral/frontopolar circuitry may need to move dorsally from Fp1 to improve tolerability. More broadly, these results establish intensity recommendations for future efficacy trials and offer a Bayesian framework for other neuromodulation dose-finding questions.

## CRediT Authorship Contribution Statement

George Kypriotakis: Conceptualization, Methodology, Software, Validation, Formal analysis, Investigation, Resources, Data curation, Writing - original draft, Writing - review & editing, Visualization, Supervision, Project administration, Funding acquisition. Francesco Versace: Conceptualization, Methodology, Software, Validation, Formal analysis, Investigation, Resources, Data curation, Writing - original draft, Writing - review & editing, Visualization, Supervision, Project administration, Funding acquisition. Lisa McTeague: Writing - review & editing. Maher Karam-Hage: Writing - review & editing. Brian A. Taylor: Writing - review & editing. Sanjay Shete: Writing - review & editing.

## Data Availability

Data are from registered clinical trials involving human participants. De-identified data and associated materials may be made available to qualified investigators upon reasonable request to the corresponding authors, subject to the scope of participant consent, institutional review and data-use agreements, privacy and confidentiality protections, and applicable NIH data-management and sharing requirements and approved study data-sharing plans.

## Declaration of Generative AI and AI-Assisted Technologies in the Writing Process

During the preparation of this work, the authors used AI-assisted tools (OpenAI ChatGPT), to help debug and improve R code used for the analyses and to support syntax and grammar corrections. The authors reviewed, verified, and edited the AI-assisted output as needed and take full responsibility for the content, analyses, and conclusions of the publication.

## Acknowledgements

We thank Alexa Baldizon, Jelena Salvador, Lauren DiPaola, and Leann Witmer for their assistance with data collection and administrative support throughout the study.

