## Supplement for "Bayesian Dose-Finding for Theta Burst Stimulation Tolerability: A Randomized Study Comparing Intermittent and Continuous Protocols at Distinct Prefrontal Targets"


George Kypriotakis, PhD[1]
Lisa McTeague PhD2, Maher Karam-Hage MD1, Brian Taylor PhD3, Sanjay Shete PhD4, Francesco Versace, PhD1

**Author Note:**

[1]The University of Texas MD Anderson Cancer Center, Department of Behavioral Science

[2] Medical University of South Carolina, Department of Psychiatry and Behavioral Sciences

[3] The University of Texas MD Anderson Cancer Center, Department of Imaging Physics

[4] The University of Texas MD Anderson Cancer Center, Department of Biostatistics

Address Correspondence to: George Kypriotakis, Ph.D., The University of Texas MD Anderson Cancer Center; P.O. Box 301407, Department of Behavioral Science Unit 1730 Houston, TX 77230-1407. Telephone: +1 713-792-5079; E-Mail:; Francesco Versace, Ph.D. The University of Texas MD Anderson Cancer Center; P.O. Box 301407, Department of Behavioral Science Unit 1730 Houston, TX 77230-1407. Telephone: +1 713 745 7933.

eMethods ... 3
- Primary endpoint definition and construction of the analytic dataset ... 3
- Dose grid, protocol coding, and sequence definition ... 3
- Binary tolerability indicators at each dose ... 3
- Primary analysis: Bayesian grouped-binomial logistic regression ... 4
  - Prior specification for the grouped primary model ... 4
- Estimation of ED70 and the dose recommendation rule ... 4
- Complementary analysis: mixed-effects logistic regression ... 5
- Curvature robustness: spline grouped models ... 5
- Nonparametric robustness: Bayesian isotonic regression ... 5
- Additional sensitivity analyses for decision thresholds and cTBS targets ... 6
- Formal protocol comparison ... 6
- Sequence effects, phenotype classification, and trajectory analyses ... 6
- Demographic subgroup analyses ... 7
- Posterior computation, diagnostics, and software ... 7
- Simulation study ... 7

Participant-Rated Symptom Data Collection and Analysis ... 9

Supplementary Tables ... 10

Supplementary Figures ... 23

### eMethods

#### Primary endpoint definition and construction of the analytic dataset

The primary tolerability endpoint was based on the final maintained stimulation intensity delivered at the end of each session rather than the highest transient intensity reached during within-session titration. For each participant i, protocol p, visit v, and session s, the session-level maintained dose was defined as the last delivered resting-motor-threshold percentage observed during that session. The participant-level maximum tolerated dose for a given protocol was then defined as the maximum of these session-final maintained doses across all available sessions for that participant-protocol combination. These Bayesian dose-finding analyses were applied to data collected under a standardized titration protocol rather than a prospective adaptive dose-assignment design, consistent with broader model-based dose-finding principles adapted from the Continual Reassessment Method and Bayesian Optimal Interval literature (O'Quigley et al., 1990; Yuan et al., 2016), and the analytic sample was determined by the number of evaluable participants available from the 2 parent cohorts rather than by a prospective dose-finding power calculation.

$$M_{ip} = \max_{s} d_{ipvs}^{(final)}$$

where $d_{ipvs}^{(final)}$ denotes the final maintained intensity in session $s$. This sustained endpoint was chosen because the scientific question for subsequent-phase trials is not whether a participant can briefly reach a peak intensity, but whether that intensity can be delivered and maintained through a full treatment session. Under the implemented titration procedure, de-escalation below the nominal 80% starting intensity was permitted and re-escalation was attempted when tolerated; therefore, observed final maintained intensities spanned 10% to 110% of resting motor threshold (RMT).

#### Dose grid, protocol coding, and sequence definition

The dose grid was derived directly from the observed titration data rather than hard-coded in advance. The script read the titration file, converted the stored proportional percent_rmt values to percentages, rounded them to whole-number percent RMT, and defined the sorted unique values as the analysis dose set $D$. The minimum and maximum observed doses, the median observed dose, and a scaling constant of 10 percentage points were then propagated throughout all model-fitting and prediction routines.
Let $d \in D$ denote a tested intensity in percent RMT. The scaled dose variable used in the parametric models was

$$x(d) = \frac{d - d_{\text{center}}}{10},$$

where $d_{\text{center}}$ is the median observed dose level. The protocol starting dose was fixed at 80% RMT, and protocol-relevant doses were defined as dose levels at or above 80% RMT.
Protocol labels were parsed directly from the stimulation field as iTBS or cTBS. Sequence was defined from the earliest recorded protocol for each participant, yielding two sequence groups: iTBS-first and cTBS-first.

#### Binary tolerability indicators at each dose

After computing each participant's protocol-specific maximum tolerated dose $M_{ip}$, the code created a binary tolerability indicator for every participant-protocol pair at every observed dose level. Specifically,

$$Y_{ip}(d) = 1\{M_{ip} \geq d\},$$

where Y indicates that participant i tolerated at least dose d under protocol p. These participant-level binary indicators were the basis for both the grouped-binomial primary analysis and the complementary mixed-effects analysis. Because each participant contributed a single maximum tolerated intensity that was then converted into binary indicators across dose thresholds, the resulting grouped counts form a cumulative distribution across dose. The grouped-binomial model therefore treats dose-specific counts as conditionally independent given the parametric dose-response curve, which is a simplifying assumption because the counts are derived from participant-level maxima. The complementary mixed-effects model relaxes this assumption by modeling participant-level binary indicators with random intercepts.$Y_{ip}(d) = 1d$

Aggregating over participants within protocol produced grouped counts $y_{pd}$ and denominators $n_{pd}$ at each dose, where

$$y_{pd} = \sum_i Y_{ip}(d), n_{pd} = \sum_i 1.$$

#### Primary analysis: Bayesian grouped-binomial logistic regression

The primary analysis fit separate grouped-binomial Bayesian logistic regression models for iTBS and cTBS as a parsimonious parametric summary of the participant-level maximum tolerated dose distribution, using a model-based dose-finding logic analogous to the Continual Reassessment Method and Bayesian Optimal Interval approaches (O'Quigley et al., 1990; Yuan et al., 2016). For protocol and dose, $pd$

$$y_{pd} \sim Binomial(n_{pd}, \pi_{pd}),$$
$$logit(\pi_{pd}) = \beta_{0p} + \beta_{1p}x(d).$$

Thus, $\pi_{pd}$ is the protocol-specific probability that a randomly selected participant would tolerate dose $d$, and $x(d)$ is dose centered at the observed median and scaled by 10 percentage points. Separate models were fit for iTBS and cTBS using a binomial likelihood with a logit link.

##### *Prior specification for the grouped primary model*

The grouped primary model used mildly informative priors chosen to encode the expected monotone decline in tolerability with increasing intensity while allowing the data to dominate inference:

$$\beta_{0p} \sim N(0, 1.5^2),$$
$$\beta_{1p} \sim N(-0.7, 0.5^2), \beta_{1p} \leq 0.$$

The intercept prior places broad prior mass over plausible tolerability values at the centering dose. The slope prior favors a modest negative dose-response and enforces the clinically reasonable restriction that tolerability should not improve with increasing intensity.

#### Estimation of ED70 and the dose recommendation rule

For each protocol, the posterior distribution of the effective dose associated with 70% tolerability (ED70) was derived from posterior draws of the grouped model. For posterior draw $s$,

$$ED70^{(s)} = d_{\text{center}} + 10\frac{logit(0.70) - \beta_0^{(s)}}{\beta_1^{(s)}}.$$

Non-finite or grossly implausible draws were excluded using a wide plausibility filter relative to the observed dose range. Posterior summaries of ED70 were then reported as means and 95% credible intervals.

Dose recommendation followed a BOIN-like posterior rule defined as the highest dose satisfying

$$Pr(\pi_{pd} \geq 0.70|\text{data}) \geq 0.80.$$

Equivalently, the recommended dose for protocol $p$ was

$$d_p = max\{d \in D: Pr(\pi_{pd} \geq 0.70|\text{data}) \geq 0.80\}.$$

If no dose satisfied this criterion, the protocol was considered not to have an acceptable recommended dose under the prespecified target. The code also generated a posterior draw-wise distribution of the recommended dose by selecting, for each posterior draw, the highest dose whose predicted tolerability exceeded the 70% target.

**Complementary analysis: mixed-effects logistic regression**

To account for within-participant dependence across doses, separate mixed-effects logistic regression models were fit for iTBS and cTBS as a complementary analysis. At the participant-dose level,

$$Y_{ip}(d) \sim Bernoulli(\pi_{ipd}),$$
$$logit(\pi_{ipd}) = \beta_{0p} + \beta_{1p}x(d) + u_i,$$

with participant-specific random intercepts

$$u_i \sim N(0, \sigma_p^2).$$

The priors for the fixed effects matched the grouped model, and the random-intercept standard deviation had a weakly informative prior:

$$\sigma_p \sim t_3(0,1), \sigma_p > 0.$$

The script distinguishes between **conditional** and **marginal** quantities. Conditional ED70 was computed from the fixed effects by setting $u_i = 0$, primarily for comparability with the grouped model. However, dose recommendations from the mixed model were based on **marginalized** tolerability probabilities that integrated over between-subject heterogeneity.

For each posterior draw $\left(\beta_0^{(s)}, \beta_1^{(s)}, \sigma^{(s)}\right)$, the code drew 200 Monte Carlo samples

$$u^{(s,r)} \sim N\left(0, {\sigma^{(s)}}^2\right), r = 1, \ldots, 200,$$

and approximated the marginal dose-specific probability as

$$\boldsymbol{\pi}_{pd}^{(s)} \approx \frac{1}{200}\sum_{r=1}^{200} logit^{-1}\left(\beta_0^{(s)} + \beta_1^{(s)}x(d) + \nu^{(s,r)}\right).$$

Posterior summaries of $\boldsymbol{\pi}_{pd}$ were then used with the same 70%/80% recommendation rule as in the primary analysis.

**Curvature robustness: spline grouped models**

To assess potential curvature misspecification, separate grouped-binomial spline models were fit for iTBS and cTBS whenever at least 4 distinct dose levels were available, following a generalized additive modeling framework (Wood, 2017). These models replaced the linear dose term with a smooth function:

$$y_{pd} \sim Binomial(n_{pd}, \pi_{pd}),$$
$$logit(\pi_{pd}) = \beta_{0p} + f_p(x(d)),$$

where f_p(d) denotes the protocol-specific penalized spline smooth of scaled dose. The basis dimension was chosen adaptively in the code as k = min(5, number of distinct dose levels - 1), with a minimum of 3. The intercept prior remained Normal(0, 1.5), and the spline standard deviation received an exponential prior. Predictions were obtained from posterior expected probabilities and the same decision rule was applied.

**Nonparametric robustness: Bayesian isotonic regression**

As a nonparametric robustness analysis, the script implemented Bayesian isotonic regression separately by protocol, consistent with order-restricted inference approaches for monotone response functions (Dykstra et al., 1991). For each dose level $d$, the dose-specific tolerability probability was assigned a Beta posterior based on observed grouped counts:

$$\pi_{pd}|y_{pd}, n_{pd} \sim Beta(1 + y_{pd}, 1 + n_{pd} - y_{pd}).$$

The code drew 5000 posterior samples by default and then imposed a monotone nonincreasing dose-response using weighted pooled-adjacent-violators regression, with weights equal to the number of participants observed at each dose. Posterior means, medians, 95% credible intervals, and

$Pr(\pi_{pd} \geq 0.70|\text{data})$ were then computed from the isotonic posterior draws. The same recommendation rule was used for the isotonic model.

**Additional sensitivity analyses for decision thresholds and cTBS targets**

The code evaluated the stability of recommendations across a grid of alternative tolerability targets and posterior decision thresholds. Specifically, target tolerability values of 0.65, 0.70, 0.75, and 0.80 were crossed with posterior probability thresholds of 0.75, 0.80, 0.85, and 0.90, and the optimal dose was recomputed under each combination using posterior expected probabilities from the grouped model.
In addition, protocol-specific cTBS sensitivity analyses examined whether lower tolerability targets of 60% or 65% would yield clinically usable recommendations when the standard 70% target was not met at protocol-relevant doses.

**Formal protocol comparison**

To compare iTBS and cTBS directly, the code fit a joint mixed-effects logistic model at the participant-dose level:

$$Y_i(d) \sim Bernoulli(\pi_i(d)),$$
$$logit(\pi_i(d)) = \beta_0 + \beta_1 x(d) + \beta_2 I(cTBS_i) + \beta_3 x(d) I(cTBS_i) + u_i,$$

with

$$u_i \sim N(0, \sigma^2).$$

Here, $\beta_2$ is the cTBS-versus-iTBS contrast at the centering dose and $\beta_3$ tests whether the dose-response slope differs by protocol. The dose slope retained a mildly informative negative prior, while the protocol main effect and interaction each received mean-zero normal priors with standard deviation 0.5. Posterior draws were used to estimate the probability that cTBS was less tolerable than iTBS overall and the probability that cTBS showed a steeper decline with increasing dose. Dose-specific protocol differences were computed from posterior expected probabilities with random effects excluded for prediction.

**Sequence effects, phenotype classification, and trajectory analyses**

Sequence effects were evaluated by fitting separate mixed-effects logistic models within iTBS and cTBS that included dose, sequence, and their interaction:

$$logit(\pi_{ipd}) = \beta_{0p} + \beta_{1p} x(d) + \beta_{2p} I(cTBS\text{-}first_i) + \beta_{3p} x(d) I(cTBS\text{-}first_i) + u_i.$$

Posterior expected probabilities were generated by dose and sequence, and posterior differences between iTBS-first and cTBS-first were summarized within each protocol.
The script also classified participants as high tolerators if their maximum tolerated dose was at least 100% RMT, moderate tolerators if it was 80%–99% RMT, and low tolerators if it was below 80% RMT. These cut points were descriptive thresholds anchored to the protocol starting dose (80% RMT) and a clinically intuitive high-intensity benchmark (100% RMT), rather than empirically derived latent classes. Within-session trajectories were summarized using the start, end, maximum, minimum, and net change in intensity and then labeled as “reached maximum,” “titrated up,” “had to decrease,” or “stayed stable.” Across-session learning was summarized by session-specific means and standard deviations of the maximum attained intensity. Among participants with both protocols, k-means clustering on the paired values was performed after standardization, with the number of clusters selected by the silhouette criterion over candidate solutions with 2 to 5 clusters.$(maxiTBS, maxcTBS)$

**Demographic subgroup analyses**

When demographic data were available, the script created age groups (<35, 35-49, and >=50 years), sex-at-birth categories, race categories, and Hispanic ethnicity labels from the demographic file and merged these participant-level data with the protocol-specific maximum tolerated dose dataset by participant identifier. Baseline demographic summaries were generated overall.

For each protocol and subgroup level, the code summarized the mean and standard deviation of the maximum tolerated intensity and the proportions achieving at least 80%, 90%, and 100% RMT, as well as the proportion classified as low tolerators (<80% RMT). For inferential subgroup comparisons, the script used Wilcoxon rank-sum tests when a subgroup variable had 2 levels within a protocol and Kruskal-Wallis tests when it had 3 or more levels. If there were fewer than 2 analyzable groups or fewer than 5 observations, the comparison was labeled insufficient. Thus, subgroup testing was nonparametric and protocol-specific, with maximum tolerated intensity as the response.

**Posterior computation, diagnostics, and software**

All Bayesian models were fit in brms using Stan-based Hamiltonian Monte Carlo (Bürkner, 2017; Carpenter et al., 2017). Under the full analysis settings in the script, each model was run with 4 chains, 4000 iterations per chain, and 1000 warmup iterations. The code used cmdstanr as the backend when available and otherwise used rstan; within-chain threading was enabled when supported by the environment. Model objects were cached to disk and refit only when the model specification changed. Model diagnostics included $\hat{R}$, bulk and tail effective sample size, absence of divergent transitions, and leave-one-out cross-validation diagnostics summarized through Pareto $k$, expected log pointwise predictive density, and standard errors. The code additionally generated posterior predictive checks and, when spline models were fit, directly compared grouped and spline models using leave-one-out cross-validation.

**Simulation study**

The appended advanced-analysis code evaluated operating characteristics of the dose-finding rule under several prespecified true dose-response scenarios. For each simulated trial, grouped binomial counts were generated at each dose level from a vector of true tolerability probabilities, and a logistic regression was then fit to the simulated grouped data. The highest dose with predicted tolerability at or above the target was selected and compared with the true optimal dose to classify each simulated trial as correct selection, overdose, underdose, or no selection.
Accordingly, the simulation component is best described as a computational operating-characteristics approximation to the Bayesian decision rule rather than a fully nested Bayesian simulation in which the full posterior model is refit inside each replicate.

**Supplementary Methods: Expanded Stimulation, Titration, and Analysis Procedures**

**Stimulation Localization and Motor Threshold Procedures**

Stimulation was delivered using a MagPro rTMS Research System with a Cool-B65 figure-of-eight coil (MagVenture A/S, Farum, Denmark). We estimated scalp positions corresponding to F3 and Fp1 using the EEG-Locator software developed by Borckardt and Hanlon. After fitting participants with a Lycra swim cap, we measured head circumference, nasion-inion distance, and intertragal distance and entered these measurements into the software. The software provided the distance and path to the selected 10-20 EEG coordinates, which were marked on the cap. Participants were then fitted with a neuronavigation tracker, and the marked scalp targets were registered in Brainsight neuronavigation software (Version 2.3.12). Resting motor threshold was estimated as the minimum stimulator intensity required to elicit a visible muscle twitch in the contralateral hand in at least 50% of trials using the TMS Motor Threshold Assessment Tool, which implements maximum-likelihood parameter estimation by sequential testing.

Once motor threshold was determined, the stimulation coil was positioned tangentially over the scalp target. For F3 stimulation, the coil handle pointed backward. For Fp1 stimulation, the coil handle pointed upward. The coil was secured with an adjustable locking arm, and the neuronavigation tracker allowed real-time monitoring and adjustment of coil position throughout stimulation.

**Titration Procedure and Endpoint Rationale**
The nominal starting intensity was 80% RMT, with upward titration in 10% increments to 90%, 100%, and 110% RMT based on participant tolerance. Per protocol, each intensity increment was scheduled at 25% intervals during the session: 90% RMT at 25% of the session, 100% RMT at 50%, and 110% RMT at 75%. The protocol permitted de-escalation below 80% RMT if participants reported the intensity as intolerable. De-escalation was implemented in 10% decrements at any point during the session, with subsequent re-escalation attempts at the following scheduled titration step as tolerated. This standardized approach yielded final maintained intensities spanning 10% to 110% RMT.

The primary tolerability endpoint was conservative by design. For each session, tolerability was defined by the final maintained intensity delivered at session completion rather than the highest transient intra-session peak. This distinction was clinically important: in the current data, 41.6% of sessions ended below their intra-session maximum, including 56.0% of cTBS sessions and 26.6% of iTBS sessions. Thus, many participants briefly reached a higher intensity but could not sustain it. The participant-level maximum tolerated dose was therefore defined as the maximum final maintained session intensity across the 3 sessions within a participant-protocol combination.

**Expanded Statistical Analysis**
The primary grouped-binomial analysis modeled the number of participants tolerating each intensity level out of the total at that level. Because each participant contributed a single maximum tolerated intensity that was converted into binary indicators across dose thresholds, the grouped-binomial model treats the resulting dose-specific counts as conditionally independent given the logistic dose-response curve. This is a simplifying assumption because the counts are derived from participant-level maxima. Priors were weakly informative: Normal(0, 1.5) for the intercept and Normal(-0.7, 0.5) with an upper bound of 0 for the slope, enforcing the physiologically plausible constraint that tolerability cannot increase with intensity. This monotonicity constraint is consistent with model-based dose-finding logic adapted from CRM and BOIN designs. Complete prior specifications, rationale, and sensitivity to alternative priors are provided in eTable 1.

Dose selection followed the prespecified rule: recommended dose = max{d : P(tolerability >=70% | data) >=80%}. This rule provides statistical confidence that the selected intensity will achieve adequate tolerability in future applications. A clinical decision lookup table for alternative target and confidence combinations is provided in eTable 2.

A complementary mixed-effects logistic regression model with participant random intercepts was fit to relax the conditional-independence assumption of the grouped model and account for within-participant dependence across dose thresholds. Recommendation probabilities from this model were marginalized over the random-effect distribution using Monte Carlo integration from posterior draws of the random-intercept standard deviation, yielding population-averaged estimates. Conditional ED70 values, with random effect set to 0, are reported for comparability with the grouped model. Additional robustness analyses included grouped spline models to relax the linearity assumption on the logit scale, Bayesian isotonic regression as a nonparametric alternative, and prior sensitivity analysis across 5 prior specifications.

Direct protocol comparison used a joint Bayesian model with a protocol-by-dose interaction term. Within-subject concordance was assessed among the 57 participants completing both protocols using Pearson correlation, intraclass correlation coefficient, Lin concordance correlation coefficient, and Bland-Altman analysis. Sequence effects were evaluated using mixed-effects models with a sequence-by-dose interaction, and carryover effects were quantified using a period-by-prior-treatment model. Session-level learning and habituation effects were assessed across the 3 within-day sessions. Demographic subgroup analyses compared maximum tolerated intensity across age group, sex at birth, race, and ethnicity using Wilcoxon rank-sum tests for 2-group comparisons and Kruskal-Wallis tests for 3 or more groups, stratified by protocol. These analyses were exploratory and not adjusted for multiple comparisons.

**Expanded Safety and Adverse Event Analysis**

Each participant-visit included one momentary assessment administered immediately after each of the 3 TMS sessions. At each assessment, participants rated headache, pain, skin or scalp irritation, facial twitching, fatigue, and fearfulness or anxiety. For each symptom, participants provided two ratings: one reflecting symptom severity during the just-completed rTMS session and one reflecting symptom severity at the time of post-session assessment. Ratings were made on an 11-point scale from 0 (not at all) to 10 (extremely). Participants could also report up to 2 additional discomfort symptoms with corresponding severity ratings.

Approximately 24 hours after each visit, participants completed the TBS screener, which assessed delayed physical symptoms, mood/activation items, and additional symptoms not captured by the structured items. Session-level total symptom burden was calculated as the sum of all immediate during-TMS symptom ratings within a session, including the 6 core symptoms and any rated other-discomfort items. Visit-level total symptom burden was calculated as the sum of session-level total burden across the 3 sessions within that participant-visit; therefore, higher values reflect both greater symptom severity and a larger number of symptoms across the visit.

Symptoms were summarized overall and by visit, session, protocol, stimulation sequence, study cohort, and demographic subgroup. A symptom was considered present if the rating was greater than 0. For immediate assessments, moderate-or-greater symptoms were defined as ratings of 4 or higher on the 0 to 10 scale, and severe symptoms were defined as ratings of 7 or higher. For 24-hour physical symptoms, moderate-or-greater symptoms were defined as ratings of 3 or higher on the 0 to 5 scale.

##### Implementation and Diagnostics

All analyses were conducted in R version 4.3 using brms for Bayesian regression and Stan for posterior sampling. Bayesian models used 4 chains of 4000 iterations, including 1000 warmup iterations. Convergence was verified using R-hat less than 1.01, effective sample size greater than 400 for bulk and tail, and absence of divergent transitions. Model fit was assessed using posterior predictive checks.

#### Supplementary Tables

##### eTable 1. Prior Sensitivity Analysis

| Prior | Intercept prior | Slope prior | iTBS (F3) recommendation | cTBS (Fp1) recommendation |
|---|---|---|---|---|
| Primary (study default) | N(0, 1.5) | N(-0.7, 0.5), ub=0 | 100% | 60% |
| Weakly informative | N(0, 3.0) | N(-0.7, 0.5), ub=0 | 100% | 60% |
| Diffuse slope | N(0, 1.5) | N(-0.5, 1.0), ub=0 | 100% | 60% |
| Skeptical (steep decline) | N(0, 1.5) | N(-1.0, 0.3), ub=0 | 100% | 60% |
| Optimistic (gentle decline) | N(0, 1.0) | N(-0.3, 0.3), ub=0 | 100% | 60% |

*Note: All priors use truncated normal for slope (upper bound = 0) to enforce monotonic decrease. Recommendations show highest dose meeting 70% tolerability with 80% confidence.*

##### eTable 2. Clinical Decision Lookup Table

| Target tolerability | Decision confidence | iTBS (F3) recommendation | cTBS (Fp1) recommendation |
|---|---|---|---|
| 65% | 75% | 100% RMT | 70% RMT |
| 70% | 75% | 100% RMT | 60% RMT |
| 75% | 75% | 90% RMT | 60% RMT |
| 80% | 75% | 90% RMT | 50% RMT |
| 65% | 80% | 100% RMT | 70% RMT |
| 70% | 80% | 100% RMT | 60% RMT |
| 75% | 80% | 90% RMT | 60% RMT |

| Target tolerability | Decision confidence | iTBS (F3) recommendation | cTBS (Fp1) recommendation |
|---|---|---|---|
| 80% | 80% | 90% RMT | 50% RMT |
| 65% | 85% | 100% RMT | 70% RMT |
| 70% | 85% | 100% RMT | 60% RMT |
| 75% | 85% | 90% RMT | 60% RMT |
| 80% | 85% | 90% RMT | 50% RMT |
| 65% | 90% | 100% RMT | 70% RMT |
| 70% | 90% | 100% RMT | 60% RMT |
| 75% | 90% | 90% RMT | 60% RMT |
| 80% | 90% | 90% RMT | 50% RMT |

*Note: Protocol starting dose: 80% RMT. Dose levels tested: 10, 20, 30, 40, 50, 60, 70, 80, 90, 100, 110% RMT.*
*Note: Highlighted row indicates protocol-specified parameters (70% tolerability, 80% confidence).*
*Note: How to use: Find your desired target tolerability and confidence level to identify the recommended starting intensity.*

### eTable 3. Within-Subject Concordance Between Protocols

| Metric | Value | 95% CI | Interpretation |
|---|---|---|---|
| Pearson r | 0.590 | (0.389, 0.737) | Moderate |
| ICC(2,1) | 0.381 | (-0.028, 0.650) | Poor agreement |
| Lin's CCC | 0.377 | (0.221, 0.513) | Not interchangeable |
| Mean difference | 22.6% RMT | - | iTBS 22.6% higher |
| Limits of agreement | -23.7 to 69.0% RMT | - | ±46.3% individual variation |

*Note: Based on 57 participants with both iTBS and cTBS data. Pearson r quantifies linear association; ICC and Lin's CCC quantify absolute agreement/concordance. ICC = Intraclass Correlation Coefficient. CCC = Concordance Correlation Coefficient.*

### eTable 4. Sequence Effect Analysis

| Protocol | Intensity (% RMT) | Mean Difference | P(iTBS-first > cTBS-first) |
|---|---|---|---|
| iTBS (F3) | 10 | 0.0% | 88.6% |
| iTBS (F3) | 20 | 0.0% | 86.7% |
| iTBS (F3) | 30 | 0.0% | 83.7% |
| iTBS (F3) | 40 | 0.0% | 77.5% |
| iTBS (F3) | 50 | 0.0% | 63.5% |
| iTBS (F3) | 60 | -0.0% | 38.0% |
| iTBS (F3) | 70 | -0.2% | 16.3% |
| iTBS (F3) | 80 | -1.1% | 8.3% |
| iTBS (F3) | 90 | -4.1% | 6.6% |
| iTBS (F3) | 100 | -12.7% | 5.9% |
| iTBS (F3) | 110 | -27.9% | 5.7% |
| cTBS (Fp1) | 10 | -0.0% | 18.9% |
| cTBS (Fp1) | 20 | -0.0% | 19.5% |
| cTBS (Fp1) | 30 | -0.0% | 21.1% |
| cTBS (Fp1) | 40 | -0.0% | 24.0% |
| cTBS (Fp1) | 50 | -0.1% | 31.8% |
| cTBS (Fp1) | 60 | -0.0% | 49.5% |
| cTBS (Fp1) | 70 | 3.3% | 68.5% |
| cTBS (Fp1) | 80 | 11.1% | 76.2% |
| cTBS (Fp1) | 90 | 8.1% | 79.6% |
| cTBS (Fp1) | 100 | 2.6% | 80.8% |
| cTBS (Fp1) | 110 | 0.6% | 81.6% |

*Note: Mean difference is P(tolerate|iTBS-first) − P(tolerate|cTBS-first). Positive values favor iTBS-first order.*

#### eTable 5. Carryover Effect Analysis

| Effect | Estimate (% RMT) | 95% CrI | P(Effect > 0) |
|---|---|---|---|
| Protocol (cTBS vs iTBS) | -16.20 | (-25.13, -7.19) | 0.0% |
| Period (2 vs 1) | 2.34 | (-9.57, 14.15) | 64.8% |
| Carryover: Prior iTBS | -3.87 | (-17.84, 9.79) | 28.9% |
| Carryover: Prior cTBS | 6.18 | (-7.66, 19.93) | 80.6% |

*Note: Model: max_tolerated ~ protocol + period + carryover_iTBS + carryover_cTBS + (1|subject). Carryover effects quantify residual influence of period 1 treatment on period 2 tolerability. Positive values indicate beneficial carryover (improved tolerability).*

#### eTable 6. Learning and Habituation Effect Across Sessions

| Protocol | Session Effect (% RMT) | P(Session Effect > 0) | Interpretation |
|---|---|---|---|
| iTBS (F3) | -0.58 (-1.66, 0.49) | 13.9% | No clear session trend |
| cTBS (Fp1) | -3.38 (-5.20, -1.53) | 0.0% | Evidence of declining tolerability across sessions |

*Note: Session effect = change in maximum tolerated intensity per additional session. Positive values indicate improvement (habituation).*

#### eTable 7. Demographic Subgroup Tolerability

| Variable | Subgroup | Protocol | N | Max Dose, mean (SD) | ≥80% RMT | ≥90% RMT | ≥100% RMT | Low Tolerator |
|---|---|---|---|---|---|---|---|---|
| Sex | Female | iTBS (F3) | 26 | 97.3 (21.3) | 80.8% | 76.9% | 69.2% | 19.2% |
| Sex | Male | iTBS (F3) | 33 | 101.5 (16.8) | 90.9% | 78.8% | 78.8% | 9.1% |
| Sex | Female | cTBS (Fp1) | 28 | 64.3 (25.2) | 32.1% | 14.3% | 14.3% | 67.9% |
| Sex | Male | cTBS (Fp1) | 34 | 88.2 (27.8) | 67.6% | 64.7% | 55.9% | 32.4% |
| Age Group | 35-49 | iTBS (F3) | 29 | 103.1 (13.4) | 93.1% | 82.8% | 79.3% | 6.9% |
| Age Group | 50+ | iTBS (F3) | 22 | 95.9 (24.6) | 77.3% | 77.3% | 72.7% | 22.7% |
| Age Group | <35 | iTBS (F3) | 8 | 97.5 (17.5) | 87.5% | 62.5% | 62.5% | 12.5% |
| Age Group | 35-49 | cTBS (Fp1) | 32 | 80.6 (26.1) | 56.2% | 40.6% | 37.5% | 43.8% |
| Age Group | 50+ | cTBS (Fp1) | 23 | 74.8 (30.3) | 47.8% | 43.5% | 34.8% | 52.2% |
| Age Group | <35 | cTBS (Fp1) | 7 | 71.4 (39.3) | 42.9% | 42.9% | 42.9% | 57.1% |
| Race | AI/AN | iTBS (F3) | 2 | 55.0 (7.1) | 0.0% | 0.0% | 0.0% | 100.0% |
| Race | Asian | iTBS (F3) | 1 | 110.0 (NA) | 100.0% | 100.0% | 100.0% | 0.0% |
| Race | Black/AA | iTBS (F3) | 34 | 98.8 (19.2) | 85.3% | 76.5% | 70.6% | 14.7% |
| Race | Multiracial | iTBS (F3) | 2 | 110.0 (0.0) | 100.0% | 100.0% | 100.0% | 0.0% |
| Race | White | iTBS (F3) | 20 | 104.0 (13.9) | 95.0% | 85.0% | 85.0% | 5.0% |
| Race | AI/AN | cTBS (Fp1) | 2 | 35.0 (7.1) | 0.0% | 0.0% | 0.0% | 100.0% |
| Race | Asian | cTBS (Fp1) | 1 | 110.0 (NA) | 100.0% | 100.0% | 100.0% | 0.0% |
| Race | Black/AA | cTBS (Fp1) | 37 | 75.1 (28.6) | 45.9% | 40.5% | 32.4% | 54.1% |
| Race | Multiracial | cTBS (Fp1) | 2 | 80.0 (42.4) | 50.0% | 50.0% | 50.0% | 50.0% |
| Race | White | cTBS (Fp1) | 20 | 84.0 (27.4) | 65.0% | 45.0% | 45.0% | 35.0% |
| Ethnicity | Hispanic | iTBS (F3) | 10 | 103.0 (12.5) | 100.0% | 80.0% | 80.0% | 0.0% |
| Ethnicity | Non-Hispanic | iTBS (F3) | 49 | 99.0 (19.9) | 83.7% | 77.6% | 73.5% | 16.3% |
| Ethnicity | Hispanic | cTBS (Fp1) | 10 | 78.0 (27.0) | 60.0% | 30.0% | 30.0% | 40.0% |
| Ethnicity | Non-Hispanic | cTBS (Fp1) | 52 | 77.3 (29.6) | 50.0% | 44.2% | 38.5% | 50.0% |

#### eTable 8. Statistical Tests for Demographic Differences

| Protocol | Variable | Test | Statistic | P-value | Significance |
|---|---|---|---|---|---|
| iTBS (F3) | Sex | Wilcoxon | 474.0000000 | 0.395 | |
| cTBS (Fp1) | Sex | Wilcoxon | 692.5000000 | 0.002 | ** |
| iTBS (F3) | Age Group | Kruskal-Wallis | 1.0691930 | 0.586 | |
| cTBS (Fp1) | Age Group | Kruskal-Wallis | 0.8341996 | 0.659 | |
| iTBS (F3) | Race | Kruskal-Wallis | 10.0612466 | 0.039 | * |
| cTBS (Fp1) | Race | Kruskal-Wallis | 7.1080819 | 0.130 | |
| iTBS (F3) | Ethnicity | Wilcoxon | 237.0000000 | 0.850 | |
| cTBS (Fp1) | Ethnicity | Wilcoxon | 253.0000000 | 0.898 | |

#### eTable 9. Baseline Participant Demographics (Expanded)

| Characteristic | Overall |
|---|---|
| N | 64 |
| Age, mean (SD) | 45.8 (8.8) |
| Age, range | 25–60 |
| Sex at birth, n (%) | |
| Male | 35 (54.7%) |
| Female | 29 (45.3%) |
| Race, n (%) | |
| Black or African American | 38 (59.4%) |
| White | 21 (32.8%) |
| Other | 5 (7.8%) |
| Ethnicity, n (%) | |
| Hispanic | 10 (15.6%) |
| Non-Hispanic | 54 (84.4%) |

#### eTable 10. Expanded Participant Flow and Protocol Completion

| Stage | Item | Value |
|---|---|---|
| Study Enrollment | Total titration observations | 2,010 |
| Study Enrollment | Unique participants | 64 |
| Study Enrollment | BRAIN study | 43 (67.2%) |
| Study Enrollment | RAPID study | 21 (32.8%) |
| Sequence Allocation | iTBS-first sequence | 33 (51.6%) |
| Sequence Allocation | cTBS-first sequence | 31 (48.4%) |
| Sequence Allocation | Completed both protocols | 57 (89.1%) |
| Session Completion | Mean sessions per participant-protocol | 2.98 |
| Session Completion | Completed all 3 sessions | 99.2% |
| Analysis - iTBS (F3) | Participants analyzed | 59 |
| Analysis - iTBS (F3) | Tolerated ≥80% RMT | 51 (86.4%) |
| Analysis - iTBS (F3) | Tolerated ≥90% RMT | 46 (78.0%) |
| Analysis - iTBS (F3) | Tolerated ≥100% RMT | 44 (74.6%) |
| Analysis - iTBS (F3) | Tolerated ≥110% RMT | 42 (71.2%) |

| Stage | Item | Value |
|---|---|---|
| Analysis - iTBS (F3) | Low tolerators (<80% RMT) | 8 (13.6%) |
| Analysis - cTBS (Fp1) | Participants analyzed | 62 |
| Analysis - cTBS (Fp1) | Tolerated ≥80% RMT | 32 (51.6%) |
| Analysis - cTBS (Fp1) | Tolerated ≥90% RMT | 26 (41.9%) |
| Analysis - cTBS (Fp1) | Tolerated ≥100% RMT | 23 (37.1%) |
| Analysis - cTBS (Fp1) | Tolerated ≥110% RMT | 22 (35.5%) |
| Analysis - cTBS (Fp1) | Low tolerators (<80% RMT) | 30 (48.4%) |
| Resting Motor Threshold | Mean (SD); Range | 55.2 (12.1); 11–85 |

*Note: Crossover design: participants could receive both iTBS and cTBS. Sequence is defined by the earliest recorded protocol (visit/session ordering). Max tolerated dose is defined as the maximum of the FINAL %RMT delivered at the end of each session (last titration step), then summarized per participant-protocol as the maximum across sessions.*

### eTable 11. Complete Posterior Estimates by Protocol, Model, and Intensity

| Protocol | Model | Intensity (% RMT) | Mean P(tolerate) | 95% CrI | P(≥70%) |
|---|---|---|---|---|---|
| iTBS (F3) | Primary | 10 | 99.6% | (99.0–99.9%) | 100.0% |
| iTBS (F3) | Primary | 20 | 99.3% | (98.5–99.8%) | 100.0% |
| iTBS (F3) | Primary | 30 | 99.0% | (97.9–99.6%) | 100.0% |
| iTBS (F3) | Primary | 40 | 98.3% | (97.0–99.2%) | 100.0% |
| iTBS (F3) | Primary | 50 | 97.3% | (95.6–98.6%) | 100.0% |
| iTBS (F3) | Primary | 60 | 95.7% | (93.6–97.4%) | 100.0% |
| iTBS (F3) | Primary | 70 | 93.1% | (90.6–95.3%) | 100.0% |
| iTBS (F3) | Primary | 80 | 89.1% | (86.0–91.9%) | 100.0% |
| iTBS (F3) | Primary | 90 | 83.3% | (79.1–87.0%) | 100.0% |
| iTBS (F3) | Primary | 100 | 75.1% | (68.8–80.7%) | 94.5% |
| iTBS (F3) | Primary | 110 | 64.5% | (55.3–73.2%) | 12.0% |
| cTBS (Fp1) | Primary | 10 | 97.7% | (96.2–98.8%) | 100.0% |
| cTBS (Fp1) | Primary | 20 | 96.3% | (94.4–97.8%) | 100.0% |
| cTBS (Fp1) | Primary | 30 | 94.1% | (91.6–96.2%) | 100.0% |
| cTBS (Fp1) | Primary | 40 | 90.7% | (87.6–93.5%) | 100.0% |
| cTBS (Fp1) | Primary | 50 | 85.7% | (82.1–89.2%) | 100.0% |
| cTBS (Fp1) | Primary | 60 | 78.6% | (74.5–82.6%) | 100.0% |
| cTBS (Fp1) | Primary | 70 | 69.1% | (64.8–73.5%) | 35.0% |
| cTBS (Fp1) | Primary | 80 | 57.8% | (52.8–62.7%) | 0.0% |
| cTBS (Fp1) | Primary | 90 | 45.5% | (39.8–51.2%) | 0.0% |
| cTBS (Fp1) | Primary | 100 | 33.8% | (27.5–40.2%) | 0.0% |
| cTBS (Fp1) | Primary | 110 | 23.8% | (17.8–30.3%) | 0.0% |
| iTBS (F3) | Sensitivity | 10 | 99.2% | (96.5–100.0%) | 100.0% |
| iTBS (F3) | Sensitivity | 20 | 98.7% | (95.0–99.9%) | 100.0% |
| iTBS (F3) | Sensitivity | 30 | 97.9% | (93.0–99.8%) | 100.0% |
| iTBS (F3) | Sensitivity | 40 | 96.6% | (90.4–99.5%) | 100.0% |
| iTBS (F3) | Sensitivity | 50 | 94.7% | (87.0–98.8%) | 100.0% |
| iTBS (F3) | Sensitivity | 60 | 91.9% | (82.8–97.4%) | 100.0% |
| iTBS (F3) | Sensitivity | 70 | 87.9% | (77.6–95.0%) | 99.8% |
| iTBS (F3) | Sensitivity | 80 | 82.7% | (71.4–91.3%) | 98.4% |
| iTBS (F3) | Sensitivity | 90 | 76.1% | (64.0–86.0%) | 85.5% |
| iTBS (F3) | Sensitivity | 100 | 68.3% | (55.7–79.6%) | 41.1% |

| Protocol | Model | Intensity (% RMT) | Mean P(tolerate) | 95% CrI | P(≥70%) |
|---|---|---|---|---|---|
| iTBS (F3) | Sensitivity | 110 | 59.5% | (46.3–72.3%) | 5.6% |
| cTBS (Fp1) | Sensitivity | 10 | 96.0% | (89.9–99.4%) | 100.0% |
| cTBS (Fp1) | Sensitivity | 20 | 93.5% | (85.8–98.3%) | 100.0% |
| cTBS (Fp1) | Sensitivity | 30 | 89.8% | (80.6–96.3%) | 100.0% |
| cTBS (Fp1) | Sensitivity | 40 | 84.7% | (74.2–92.9%) | 99.7% |
| cTBS (Fp1) | Sensitivity | 50 | 78.1% | (66.8–87.6%) | 92.7% |
| cTBS (Fp1) | Sensitivity | 60 | 70.0% | (58.3–80.5%) | 50.8% |
| cTBS (Fp1) | Sensitivity | 70 | 60.6% | (49.2–71.7%) | 5.1% |
| cTBS (Fp1) | Sensitivity | 80 | 50.6% | (39.5–61.8%) | 0.0% |
| cTBS (Fp1) | Sensitivity | 90 | 40.5% | (29.9–51.5%) | 0.0% |
| cTBS (Fp1) | Sensitivity | 100 | 31.0% | (21.1–41.8%) | 0.0% |
| cTBS (Fp1) | Sensitivity | 110 | 22.7% | (13.7–32.9%) | 0.0% |

*Note: Abbreviations: CrI, credible interval; P(≥70%), posterior probability that true tolerability exceeds 70%.*

#### eTable 12. Expected Tolerators per 100 Patients by Intensity

| Protocol | Intensity (% RMT) | Expected per 100 (95% CrI) | P(≥70%) |
|---|---|---|---|
| iTBS (F3) | 10 | 100 (99–100) | 100.0% |
| iTBS (F3) | 20 | 99 (99–100) | 100.0% |
| iTBS (F3) | 30 | 99 (98–100) | 100.0% |
| iTBS (F3) | 40 | 98 (97–99) | 100.0% |
| iTBS (F3) | 50 | 97 (96–99) | 100.0% |
| iTBS (F3) | 60 | 96 (94–97) | 100.0% |
| iTBS (F3) | 70 | 93 (91–95) | 100.0% |
| iTBS (F3) | 80 | 89 (86–92) | 100.0% |
| iTBS (F3) | 90 | 83 (79–87) | 100.0% |
| iTBS (F3) | 100 | 75 (69–81) | 94.5% |
| iTBS (F3) | 110 | 65 (55–73) | 12.0% |
| cTBS (Fp1) | 10 | 98 (96–99) | 100.0% |
| cTBS (Fp1) | 20 | 96 (94–98) | 100.0% |
| cTBS (Fp1) | 30 | 94 (92–96) | 100.0% |
| cTBS (Fp1) | 40 | 91 (88–94) | 100.0% |
| cTBS (Fp1) | 50 | 86 (82–89) | 100.0% |
| cTBS (Fp1) | 60 | 79 (75–83) | 100.0% |
| cTBS (Fp1) | 70 | 69 (65–74) | 35.0% |
| cTBS (Fp1) | 80 | 58 (53–63) | 0.0% |
| cTBS (Fp1) | 90 | 45 (40–51) | 0.0% |
| cTBS (Fp1) | 100 | 34 (28–40) | 0.0% |
| cTBS (Fp1) | 110 | 24 (18–30) | 0.0% |

#### eTable 13. Formal Protocol Comparison at Each Intensity

| Intensity (% RMT) | iTBS P(tolerate) | cTBS P(tolerate) | Difference (iTBS − cTBS) | P(iTBS > cTBS) | P(Diff > 10%) |
|---|---|---|---|---|---|
| 10 | 99.6% (99.0–99.9) | 97.7% (96.3–98.8) | 1.9% (0.7–3.4) | 99.9% | 0.0% |
| 20 | 99.4% (98.6–99.8) | 96.3% (94.4–97.8) | 3.0% (1.3–5.0) | 100.0% | 0.0% |
| 30 | 99.0% (97.9–99.6) | 94.2% (91.7–96.2) | 4.8% (2.5–7.4) | 100.0% | 0.0% |
| 40 | 98.3% (96.9–99.3) | 90.9% (87.8–93.5) | 7.5% (4.5–10.7) | 100.0% | 6.0% |

| Intensity (% RMT) | iTBS P(tolerate) | cTBS P(tolerate) | Difference (iTBS − cTBS) | P(iTBS > cTBS) | P(Diff > 10%) |
|---|---|---|---|---|---|
| 50 | 97.3% (95.6–98.6) | 85.9% (82.2–89.2) | 11.5% (7.7–15.3) | 100.0% | 76.9% |
| 60 | 95.7% (93.5–97.4) | 78.8% (74.8–82.6) | 16.9% (12.6–21.3) | 100.0% | 99.9% |
| 70 | 93.1% (90.5–95.4) | 69.4% (65.0–73.6) | 23.8% (18.9–28.6) | 100.0% | 100.0% |
| 80 | 89.2% (86.0–92.0) | 58.0% (53.1–62.7) | 31.2% (25.6–36.8) | 100.0% | 100.0% |
| 90 | 83.3% (79.1–87.1) | 45.7% (40.1–51.2) | 37.6% (30.8–44.4) | 100.0% | 100.0% |
| 100 | 75.1% (69.0–80.8) | 33.9% (27.9–40.2) | 41.2% (32.4–49.6) | 100.0% | 100.0% |
| 110 | 64.5% (55.4–73.2) | 23.9% (18.1–30.3) | 40.6% (29.4–51.2) | 100.0% | 100.0% |

*Note: Joint model: tolerated | trials(n) ~ dose_scaled * protocol. Difference = iTBS minus cTBS tolerability probability. P(iTBS > cTBS) = posterior probability iTBS has higher tolerability at that dose.*

#### eTable 14. Tolerability Phenotype Characteristics

| Protocol | Phenotype | N (%) | Mean +/- SD | Range |
|---|---|---|---|---|
| iTBS (F3) | High tolerator (≥100%) | 44 (74.6%) | 109.5 ± 2.1 | 100–110 |
| iTBS (F3) | Moderate tolerator (80-99%) | 7 (11.9%) | 82.9 ± 4.9 | 80–90 |
| iTBS (F3) | Low tolerator (<80%) | 8 (13.6%) | 60.0 ± 14.1 | 30–70 |
| cTBS (Fp1) | High tolerator (≥100%) | 23 (37.1%) | 109.6 ± 2.1 | 100–110 |
| cTBS (Fp1) | Moderate tolerator (80-99%) | 9 (14.5%) | 83.3 ± 5.0 | 80–90 |
| cTBS (Fp1) | Low tolerator (<80%) | 30 (48.4%) | 51.0 ± 14.5 | 20–70 |

*Note: Values are maximum tolerated intensity (% RMT). Phenotype classification based on highest intensity achieved.*

#### eTable 15. Immediate Core Symptoms by Protocol and Timepoint

| Protocol | Symptom | Timepoint | Observations, No. | Any symptom, No. (%) | Moderate-or-greater, No. (%) | Severe, No. (%) | Mean rating | Mean if present | Maximum rating |
|---|---|---|---|---|---|---|---|---|---|
| iTBS (F3) | Facial twitching | During TMS | 177 | 118/177 (66.7%) | 65/177 (36.7%) | 32/177 (18.1%) | 3.12 | 4.68 | 10 |
| iTBS (F3) | Skin/scalp irritation | During TMS | 177 | 89/177 (50.3%) | 49/177 (27.7%) | 23/177 (13.0%) | 2.19 | 4.35 | 10 |
| iTBS (F3) | Pain | During TMS | 177 | 87/177 (49.2%) | 36/177 (20.3%) | 12/177 (6.8%) | 1.79 | 3.64 | 10 |
| iTBS (F3) | Headache | During TMS | 177 | 58/177 (32.8%) | 29/177 (16.4%) | 13/177 (7.3%) | 1.27 | 3.88 | 8 |
| iTBS (F3) | Fearfulness/anxiety | During TMS | 177 | 47/177 (26.6%) | 21/177 (11.9%) | 5/177 (2.8%) | 0.91 | 3.43 | 9 |
| iTBS (F3) | Fatigue | During TMS | 177 | 22/177 (12.4%) | 11/177 (6.2%) | 3/177 (1.7%) | 0.46 | 3.68 | 10 |
| iTBS (F3) | Headache | Post-session | 177 | 42/177 (23.7%) | 16/177 (9.0%) | 6/177 (3.4%) | 0.72 | 3.05 | 8 |
| iTBS (F3) | Pain | Post-session | 177 | 28/177 (15.8%) | 2/177 (1.1%) | 0/177 (0.0%) | 0.29 | 1.82 | 4 |
| iTBS (F3) | Fatigue | Post-session | 177 | 21/177 (11.9%) | 8/177 (4.5%) | 1/177 (0.6%) | 0.35 | 2.95 | 7 |
| iTBS (F3) | Skin/scalp irritation | Post-session | 176 | 15/176 (8.5%) | 1/176 (0.6%) | 0/176 (0.0%) | 0.15 | 1.73 | 4 |
| iTBS (F3) | Fearfulness/anxiety | Post-session | 177 | 10/177 (5.6%) | 2/177 (1.1%) | 0/177 (0.0%) | 0.11 | 1.90 | 4 |
| iTBS (F3) | Facial twitching | Post-session | 177 | 6/177 (3.4%) | 0/177 (0.0%) | 0/177 (0.0%) | 0.06 | 1.67 | 2 |
| cTBS (Fp1) | Pain | During TMS | 183 | 140/183 (76.5%) | 98/183 (53.6%) | 42/183 (23.0%) | 3.97 | 5.19 | 10 |

| Protocol | Symptom | Timepoint | Observations, No. | Any symptom, No. (%) | Moderate-or-greater, No. (%) | Severe, No. (%) | Mean rating | Mean if present | Maximum rating |
|---|---|---|---|---|---|---|---|---|---|
| cTBS (Fp1) | Facial twitching | During TMS | 183 | 124/183 (67.8%) | 87/183 (47.5%) | 42/183 (23.0%) | 3.69 | 5.44 | 10 |
| cTBS (Fp1) | Skin/scalp irritation | During TMS | 183 | 116/183 (63.4%) | 89/183 (48.6%) | 56/183 (30.6%) | 3.81 | 6.02 | 10 |
| cTBS (Fp1) | Fearfulness/anxiety | During TMS | 183 | 72/183 (39.3%) | 42/183 (23.0%) | 23/183 (12.6%) | 1.97 | 5.01 | 10 |
| cTBS (Fp1) | Headache | During TMS | 183 | 71/183 (38.8%) | 55/183 (30.1%) | 28/183 (15.3%) | 2.17 | 5.61 | 10 |
| cTBS (Fp1) | Fatigue | During TMS | 183 | 21/183 (11.5%) | 10/183 (5.5%) | 3/183 (1.6%) | 0.50 | 4.33 | 10 |
| cTBS (Fp1) | Headache | Post-session | 183 | 43/183 (23.5%) | 14/183 (7.7%) | 1/183 (0.5%) | 0.68 | 2.91 | 7 |
| cTBS (Fp1) | Pain | Post-session | 183 | 43/183 (23.5%) | 7/183 (3.8%) | 0/183 (0.0%) | 0.50 | 2.14 | 5 |
| cTBS (Fp1) | Skin/scalp irritation | Post-session | 183 | 28/183 (15.3%) | 1/183 (0.5%) | 0/183 (0.0%) | 0.31 | 2.00 | 4 |
| cTBS (Fp1) | Fearfulness/anxiety | Post-session | 183 | 24/183 (13.1%) | 8/183 (4.4%) | 1/183 (0.5%) | 0.34 | 2.62 | 7 |
| cTBS (Fp1) | Fatigue | Post-session | 183 | 22/183 (12.0%) | 7/183 (3.8%) | 1/183 (0.5%) | 0.35 | 2.91 | 7 |
| cTBS (Fp1) | Facial twitching | Post-session | 183 | 20/183 (10.9%) | 6/183 (3.3%) | 1/183 (0.5%) | 0.27 | 2.50 | 10 |

#### eTable 16. Visit-Level Immediate Symptom Burden by Protocol

| Protocol | Visits, No. | Participants, No. | Mean visit total burden | SD | Median | Mean session burden | Any moderate-or-greater visit, % | Any severe rating, % | Mean maximum rating |
|---|---|---|---|---|---|---|---|---|---|
| iTBS (F3) | 59 | 59 | 32.1 | 27.9 | 27.0 | 10.7 | 69.5% | 39.0% | 5.5 |
| cTBS (Fp1) | 61 | 61 | 52.6 | 38.5 | 45.0 | 17.5 | 90.2% | 55.7% | 6.8 |

#### eTable 17. Paired Within-Participant Immediate Symptom Burden Comparisons by Protocol

| Metric | Paired participants, No. | iTBS mean | cTBS mean | Mean difference, cTBS - iTBS | Median difference, cTBS - iTBS | P value |
|---|---|---|---|---|---|---|
| Visit-level total burden | 57 | 32.25 | 51.49 | 19.25 | 16.00 | <.001 |
| Visit-level core burden | 57 | 29.23 | 47.42 | 18.19 | 15.00 | <.001 |
| Mean session burden | 57 | 10.75 | 17.16 | 6.42 | 5.33 | <.001 |
| Maximum single-session rating | 57 | 5.49 | 6.75 | 1.26 | 1.00 | <.001 |
| Moderate-or-greater ratings per visit | 57 | 3.96 | 6.63 | 2.67 | 3.00 | <.001 |
| Any moderate-or-greater symptom | 57 | 0.70 | 0.89 | 0.19 | 0.00 | .008 |
| Any severe rating | 57 | 0.39 | 0.56 | 0.18 | 0.00 | .020 |

#### eTable 18. Mixed Model Results for Immediate Symptom Burden and Moderate-or-Greater Symptoms

| Model | Term | Estimate (95% CI) | SE | Statistic | P value | OR (95% CI) |
|---|---|---|---|---|---|---|
| Linear mixed model: total immediate symptom burden | (Intercept) | 14.77 (11.79 to 17.75) | 1.52 | 9.72 | <.001 | |
| Linear mixed model: total immediate symptom burden | protocolcTBS | 4.80 (2.60 to 7.00) | 1.12 | 4.28 | <.001 | |
| Linear mixed model: total immediate symptom burden | final_pct_sc | -2.44 (-4.81 to -0.08) | 1.21 | -2.02 | .044 | |
| Linear mixed model: total immediate symptom burden | session_factor2 | -2.03 (-3.95 to -0.12) | 0.98 | -2.08 | .039 | |
| Linear mixed model: total immediate symptom burden | session_factor3 | -3.39 (-5.31 to -1.47) | 0.98 | -3.47 | <.001 | |
| Linear mixed model: total immediate symptom burden | visit_factor2 | -2.11 (-3.74 to -0.48) | 0.83 | -2.54 | .012 | |
| Linear mixed model: total immediate symptom burden | protocolcTBS:final_pct_sc | 0.58 (-1.56 to 2.72) | 1.09 | 0.53 | .596 | |
| Mixed logistic model: any moderate-or-greater symptom | (Intercept) | 2.42 (1.31 to 3.53) | 0.57 | 4.27 | <.001 | 11.21 (3.70 to 34.00) |
| Mixed logistic model: any moderate-or-greater symptom | protocolcTBS | 1.71 (0.72 to 2.71) | 0.51 | 3.38 | <.001 | 5.55 (2.05 to 15.02) |
| Mixed logistic model: any moderate-or-greater symptom | final_pct_sc | -0.82 (-1.74 to 0.10) | 0.47 | -1.74 | .082 | 0.44 (0.18 to 1.11) |
| Mixed logistic model: any moderate-or-greater symptom | session_factor2 | -1.51 (-2.32 to -0.69) | 0.42 | -3.61 | <.001 | 0.22 (0.10 to 0.50) |
| Mixed logistic model: any moderate-or-greater symptom | session_factor3 | -1.68 (-2.50 to -0.85) | 0.42 | -3.99 | <.001 | 0.19 (0.08 to 0.43) |
| Mixed logistic model: any moderate-or-greater symptom | visit_factor2 | -1.13 (-1.88 to -0.39) | 0.38 | -2.98 | .003 | 0.32 (0.15 to 0.68) |
| Mixed logistic model: any moderate-or-greater symptom | protocolcTBS:final_pct_sc | 0.69 (-0.21 to 1.58) | 0.46 | 1.50 | .133 | 1.99 (0.81 to 4.87) |
| Linear mixed model: protocol by sex | (Intercept) | 11.94 (8.00 to 15.88) | 2.01 | 5.94 | <.001 | |
| Linear mixed model: protocol by sex | protocolcTBS | 4.45 (1.25 to 7.64) | 1.63 | 2.73 | .007 | |

| Model | Term | Estimate (95% CI) | SE | Statistic | P value | OR (95% CI) |
|---|---|---|---|---|---|---|
| Linear mixed model: protocol by sex | sex_labelMale | 4.98 (-0.00 to 9.96) | 2.54 | 1.96 | .055 | |
| Linear mixed model: protocol by sex | final_pct_sc | -2.30 (-3.95 to -0.64) | 0.84 | -2.72 | .007 | |
| Linear mixed model: protocol by sex | session_factor2 | -2.02 (-3.94 to -0.10) | 0.98 | -2.06 | .040 | |
| Linear mixed model: protocol by sex | session_factor3 | -3.36 (-5.28 to -1.44) | 0.98 | -3.44 | <.001 | |
| Linear mixed model: protocol by sex | visit_factor2 | -2.13 (-3.76 to -0.50) | 0.83 | -2.57 | .011 | |
| Linear mixed model: protocol by sex | protocolcTBS:sex_labelMale | 0.42 (-3.00 to 3.84) | 1.74 | 0.24 | .810 | |
| Mixed logistic model: protocol by sex | (Intercept) | 1.84 (0.47 to 3.20) | 0.70 | 2.64 | .008 | 6.28 (1.60 to 24.63) |
| Mixed logistic model: protocol by sex | protocolcTBS | 1.59 (0.19 to 3.00) | 0.72 | 2.22 | .026 | 4.92 (1.21 to 20.06) |
| Mixed logistic model: protocol by sex | sex_labelMale | 0.63 (-1.03 to 2.30) | 0.85 | 0.75 | .456 | 1.89 (0.36 to 9.99) |
| Mixed logistic model: protocol by sex | final_pct_sc | -0.42 (-1.12 to 0.28) | 0.36 | -1.18 | .237 | 0.66 (0.33 to 1.32) |
| Mixed logistic model: protocol by sex | session_factor2 | -1.48 (-2.29 to -0.67) | 0.42 | -3.56 | <.001 | 0.23 (0.10 to 0.51) |
| Mixed logistic model: protocol by sex | session_factor3 | -1.63 (-2.45 to -0.82) | 0.42 | -3.92 | <.001 | 0.20 (0.09 to 0.44) |
| Mixed logistic model: protocol by sex | visit_factor2 | -1.14 (-1.88 to -0.40) | 0.38 | -3.01 | .003 | 0.32 (0.15 to 0.67) |
| Mixed logistic model: protocol by sex | protocolcTBS:sex_labelMale | 0.40 (-1.15 to 1.94) | 0.79 | 0.50 | .615 | 1.49 (0.32 to 6.96) |

#### eTable 19. Within-Visit Immediate Symptom Burden Trajectory by Protocol and Session

| Protocol | Session | Sessions, No. | Participants, No. | Mean total burden (SE) | Mean core burden | Any symptom, % | Any moderate-or-greater, % | Mean maximum rating |
|---|---|---|---|---|---|---|---|---|
| iTBS (F3) | 1 | 59 | 59 | 12.9 (1.4) | 11.8 | 89.8% | 62.7% | 4.9 |
| iTBS (F3) | 2 | 59 | 59 | 10.0 (1.3) | 9.1 | 86.4% | 47.5% | 4.1 |
| iTBS (F3) | 3 | 59 | 59 | 9.2 (1.2) | 8.4 | 83.1% | 47.5% | 3.9 |
| cTBS (Fp1) | 1 | 61 | 61 | 19.1 (1.7) | 17.7 | 96.7% | 86.9% | 6.5 |
| cTBS (Fp1) | 2 | 61 | 61 | 17.7 (1.8) | 16.2 | 90.2% | 72.1% | 5.5 |
| cTBS (Fp1) | 3 | 61 | 61 | 15.8 (1.8) | 14.5 | 86.9% | 68.9% | 4.9 |

#### eTable 20. Immediate Post-Session Resolution Among Symptoms Present During TMS

| Protocol | Symptom | Present during TMS, No. | Resolved, No. (%) | Improved but not resolved, No. (%) | Persistent post-session, No. (%) | Worsened, No. (%) | Mean change, post - during |
|---|---|---|---|---|---|---|---|
| iTBS (F3) | Facial twitching | 118 | 112/118 (94.9%) | 6/118 (5.1%) | 6/118 (5.1%) | 0/118 (0.0%) | -4.59 |
| iTBS (F3) | Skin/scalp irritation | 88 | 76/88 (86.4%) | 10/88 (11.4%) | 12/88 (13.6%) | 0/88 (0.0%) | -4.07 |
| iTBS (F3) | Pain | 87 | 62/87 (71.3%) | 22/87 (25.3%) | 25/87 (28.7%) | 0/87 (0.0%) | -3.10 |
| iTBS (F3) | Headache | 58 | 23/58 (39.7%) | 20/58 (34.5%) | 35/58 (60.3%) | 3/58 (5.2%) | -1.95 |
| iTBS (F3) | Fearfulness/anxiety | 47 | 37/47 (78.7%) | 8/47 (17.0%) | 10/47 (21.3%) | 0/47 (0.0%) | -3.02 |
| iTBS (F3) | Fatigue | 22 | 4/22 (18.2%) | 6/22 (27.3%) | 18/22 (81.8%) | 0/22 (0.0%) | -1.18 |
| cTBS (Fp1) | Pain | 140 | 99/140 (70.7%) | 39/140 (27.9%) | 41/140 (29.3%) | 0/140 (0.0%) | -4.55 |
| cTBS (Fp1) | Facial twitching | 124 | 106/124 (85.5%) | 15/124 (12.1%) | 18/124 (14.5%) | 0/124 (0.0%) | -5.08 |
| cTBS (Fp1) | Skin/scalp irritation | 116 | 89/116 (76.7%) | 25/116 (21.6%) | 27/116 (23.3%) | 0/116 (0.0%) | -5.54 |
| cTBS (Fp1) | Fearfulness/anxiety | 72 | 48/72 (66.7%) | 18/72 (25.0%) | 24/72 (33.3%) | 0/72 (0.0%) | -4.14 |
| cTBS (Fp1) | Headache | 71 | 35/71 (49.3%) | 28/71 (39.4%) | 36/71 (50.7%) | 2/71 (2.8%) | -4.11 |
| cTBS (Fp1) | Fatigue | 21 | 6/21 (28.6%) | 10/21 (47.6%) | 15/21 (71.4%) | 0/21 (0.0%) | -2.14 |

#### eTable 21. 24-Hour TBS Screener Physical Symptoms by Protocol

| Protocol | Symptom | Visits, No. | Any symptom, No. (%) | Moderate-or-greater, No. (%) | Mean rating | Mean if present | Maximum rating |
|---|---|---|---|---|---|---|---|
| iTBS (F3) | Headache | 59 | 12/59 (20.3%) | 4/59 (6.8%) | 0.44 | 2.17 | 5 |
| iTBS (F3) | Appetite loss | 59 | 3/59 (5.1%) | 3/59 (5.1%) | 0.20 | 4.00 | 5 |
| iTBS (F3) | Weakness | 59 | 3/59 (5.1%) | 3/59 (5.1%) | 0.19 | 3.67 | 5 |
| iTBS (F3) | Scalp pain | 59 | 2/59 (3.4%) | 0/59 (0.0%) | 0.03 | 1.00 | 1 |
| iTBS (F3) | Skin changes | 59 | 2/59 (3.4%) | 0/59 (0.0%) | 0.05 | 1.50 | 2 |
| iTBS (F3) | Vision changes | 59 | 2/59 (3.4%) | 2/59 (3.4%) | 0.14 | 4.00 | 5 |
| iTBS (F3) | Arm/hand pain | 59 | 1/59 (1.7%) | 0/59 (0.0%) | 0.02 | 1.00 | 1 |
| iTBS (F3) | Ear ringing | 59 | 1/59 (1.7%) | 1/59 (1.7%) | 0.05 | 3.00 | 3 |
| iTBS (F3) | Hearing changes | 59 | 0/59 (0.0%) | 0/59 (0.0%) | 0.00 | | 0 |
| iTBS (F3) | Loss of dexterity | 59 | 0/59 (0.0%) | 0/59 (0.0%) | 0.00 | | 0 |
| iTBS (F3) | Nausea/vomiting | 59 | 0/59 (0.0%) | 0/59 (0.0%) | 0.00 | | 0 |
| iTBS (F3) | Other pain | 59 | 0/59 (0.0%) | 0/59 (0.0%) | 0.00 | | 0 |
| iTBS (F3) | Rash | 59 | 0/59 (0.0%) | 0/59 (0.0%) | 0.00 | | 0 |
| cTBS (Fp1) | Headache | 61 | 11/61 (18.0%) | 2/61 (3.3%) | 0.34 | 1.91 | 5 |
| cTBS (Fp1) | Weakness | 61 | 8/61 (13.1%) | 2/61 (3.3%) | 0.26 | 2.00 | 5 |
| cTBS (Fp1) | Arm/hand pain | 61 | 5/61 (8.2%) | 2/61 (3.3%) | 0.21 | 2.60 | 4 |
| cTBS (Fp1) | Scalp pain | 61 | 4/61 (6.6%) | 2/61 (3.3%) | 0.18 | 2.75 | 5 |
| cTBS (Fp1) | Appetite loss | 61 | 3/61 (4.9%) | 2/61 (3.3%) | 0.18 | 3.67 | 5 |
| cTBS (Fp1) | Ear ringing | 61 | 3/61 (4.9%) | 0/61 (0.0%) | 0.07 | 1.33 | 2 |
| cTBS (Fp1) | Loss of dexterity | 61 | 3/61 (4.9%) | 1/61 (1.6%) | 0.11 | 2.33 | 4 |
| cTBS (Fp1) | Hearing changes | 61 | 2/61 (3.3%) | 2/61 (3.3%) | 0.11 | 3.50 | 4 |
| cTBS (Fp1) | Nausea/vomiting | 61 | 2/61 (3.3%) | 1/61 (1.6%) | 0.08 | 2.50 | 4 |
| cTBS (Fp1) | Other pain | 61 | 1/61 (1.6%) | 0/61 (0.0%) | 0.02 | 1.00 | 1 |
| cTBS (Fp1) | Skin changes | 61 | 1/61 (1.6%) | 0/61 (0.0%) | 0.02 | 1.00 | 1 |

| Protocol | Symptom | Visits, No. | Any symptom, No. (%) | Moderate-or-greater, No. (%) | Mean rating | Mean if present | Maximum rating |
|---|---|---|---|---|---|---|---|
| cTBS (Fp1) | Vision changes | 61 | 1/61 (1.6%) | 1/61 (1.6%) | 0.08 | 5.00 | 5 |
| cTBS (Fp1) | Rash | 61 | 0/61 (0.0%) | 0/61 (0.0%) | 0.00 | | 0 |

*Note: This supplementary table includes all structured physical symptoms, including symptoms with 0 reported prevalence in a protocol.*

#### eTable 22. 24-Hour Mood and Activation Items by Protocol

| Protocol | Item | Visits, No. | Yes, No. (%) |
|---|---|---|---|
| iTBS (F3) | Unusual energy/motivation | 59 | 7/59 (11.9%) |
| iTBS (F3) | Unusually good/high | 59 | 4/59 (6.8%) |
| iTBS (F3) | Unusually irritable | 58 | 2/58 (3.4%) |
| cTBS (Fp1) | Unusual energy/motivation | 61 | 6/61 (9.8%) |
| cTBS (Fp1) | Unusually good/high | 61 | 6/61 (9.8%) |
| cTBS (Fp1) | Unusually irritable | 61 | 6/61 (9.8%) |

#### eTable 23. Optional Other-Discomfort Reports by Protocol and Timepoint

| Protocol | Timepoint | Sessions, No. | Sessions with other, No. (%) | Mean other burden | Mean maximum other rating | Maximum other rating |
|---|---|---|---|---|---|---|
| iTBS (F3) | During TMS | 177 | 41/177 (23.2%) | 0.97 | 0.95 | 9 |
| iTBS (F3) | Post-session | 177 | 41/177 (23.2%) | 0.15 | 0.15 | 8 |
| cTBS (Fp1) | During TMS | 183 | 52/183 (28.4%) | 1.42 | 1.23 | 10 |
| cTBS (Fp1) | Post-session | 183 | 52/183 (28.4%) | 0.31 | 0.25 | 10 |

#### eTable 24. Other-Discomfort Free-Text Reports During TMS

| Protocol | Free-text category | Free-text entry | Entries, No. | Participants, No. | Mean rating | Maximum rating |
|---|---|---|---|---|---|---|
| iTBS (F3) | Teeth/jaw sensation | teeth chattering | 9 | 3 | 4.33 | 9 |
| iTBS (F3) | Nasal/sinus | sinus muscles contracting | 3 | 1 | 2.00 | 3 |
| iTBS (F3) | Pain/tapping sensation | irritated by the tapping sensation, but was not painful | 3 | 1 | 6.00 | 7 |
| iTBS (F3) | Teeth/jaw sensation | jaw twitching, which caused teeth clattering | 3 | 1 | 3.67 | 4 |
| iTBS (F3) | Teeth/jaw sensation | left jaw clenching | 3 | 1 | 8.00 | 8 |
| iTBS (F3) | Eye/watering | teary eyed | 2 | 1 | 0.00 | 0 |
| iTBS (F3) | Other | facial tightening | 2 | 1 | 3.00 | 4 |
| iTBS (F3) | Pain/tapping sensation | tapping sensation | 2 | 1 | 2.00 | 3 |
| iTBS (F3) | Arm/hand sensation | right arm pain, pulled muscle, perhaps due to neck pillow | 1 | 1 | 8.00 | 8 |
| iTBS (F3) | Eye/watering | corner of eye twitching/discomfort ; first stimulation after each pause, there was a pinching sensation near the corner of his left eye | 1 | 1 | 3.00 | 3 |
| iTBS (F3) | Eye/watering | eyeball vibrating | 1 | 1 | 6.00 | 6 |

| Protocol | Free-text category | Free-text entry | Entries, No. | Participants, No. | Mean rating | Maximum rating |
|---|---|---|---|---|---|---|
| iTBS (F3) | Eye/watering | feeling of eye/back of cornea vibrating | 1 | 1 | 7.00 | 7 |
| iTBS (F3) | Eye/watering | teary eyed (right eye) (as a result of sensation in right temple) | 1 | 1 | 1.00 | 1 |
| iTBS (F3) | Nasal/sinus | Nasal irritation | 1 | 1 | 3.00 | 3 |
| iTBS (F3) | Pain/tapping sensation | tapping feeling head | 1 | 1 | 3.00 | 3 |
| iTBS (F3) | Pain/tapping sensation | tapping sensation more towards the back of the head | 1 | 1 | 7.00 | 7 |
| iTBS (F3) | Sound irritation | irritated by sound | 1 | 1 | 5.00 | 5 |
| iTBS (F3) | Sound irritation | participant finds the sound to be irritating | 1 | 1 | 5.00 | 5 |
| iTBS (F3) | Teeth/jaw sensation | Teeth tingling feeling | 1 | 1 | 2.00 | 2 |
| iTBS (F3) | Teeth/jaw sensation | bit tongue while teeth were chattering | 1 | 1 | 4.00 | 4 |
| iTBS (F3) | Teeth/jaw sensation | left side of jaw/teeth chattering during stimulation | 1 | 1 | 6.00 | 6 |
| iTBS (F3) | Teeth/jaw sensation | teeth clattering | 1 | 1 | 1.00 | 1 |
| iTBS (F3) | Vision change | Vision blurred | 1 | 1 | 3.00 | 3 |
| cTBS (Fp1) | Eye/watering | eye pain | 4 | 2 | 5.00 | 10 |
| cTBS (Fp1) | Eye/watering | feeling of twitching between left eye and nose | 3 | 1 | 7.00 | 8 |
| cTBS (Fp1) | Eye/watering | watering eyes | 3 | 2 | 6.67 | 10 |
| cTBS (Fp1) | Pain/tapping sensation | painful tapping on head | 3 | 1 | 4.00 | 5 |
| cTBS (Fp1) | Teeth/jaw sensation | teeth chattering | 3 | 2 | 3.33 | 5 |
| cTBS (Fp1) | Ear ringing/tinnitus | tinnitus | 2 | 1 | 0.00 | 0 |
| cTBS (Fp1) | Eye/watering | Watering eyes | 2 | 1 | 5.50 | 7 |
| cTBS (Fp1) | Eye/watering | eye fatigue | 2 | 1 | 0.00 | 0 |
| cTBS (Fp1) | Eye/watering | sinus pressure caused teary eyes | 2 | 1 | 5.50 | 6 |
| cTBS (Fp1) | Eye/watering | watering eye | 2 | 1 | 6.50 | 7 |
| cTBS (Fp1) | Eye/watering | watery eyes | 2 | 1 | 1.00 | 2 |
| cTBS (Fp1) | Nasal/sinus | Nasal Discomfort | 2 | 1 | 3.50 | 4 |
| cTBS (Fp1) | Other | Agitation | 2 | 1 | 3.00 | 4 |
| cTBS (Fp1) | Teeth/jaw sensation | teeth pain | 2 | 1 | 6.00 | 7 |
| cTBS (Fp1) | Arm/hand sensation | arm twitching | 1 | 1 | 0.00 | 0 |
| cTBS (Fp1) | Eye/watering | Eye pain | 1 | 1 | 1.00 | 1 |
| cTBS (Fp1) | Eye/watering | Watering eyes (recorded for 1st MA) | 1 | 1 | 0.00 | 0 |
| cTBS (Fp1) | Eye/watering | eye fatigue (and twitching of left eye) | 1 | 1 | 5.00 | 5 |

| Protocol | Free-text category | Free-text entry | Entries, No. | Participants, No. | Mean rating | Maximum rating |
|---|---|---|---|---|---|---|
| cTBS (Fp1) | Eye/watering | left eye pain (pain in the back of the eye) (going away at the moment) | 1 | 1 | 2.00 | 2 |
| cTBS (Fp1) | Eye/watering | sinus pressure caused eye to water | 1 | 1 | 6.00 | 6 |
| cTBS (Fp1) | Eye/watering | teary eyed | 1 | 1 | 0.00 | 0 |
| cTBS (Fp1) | Eye/watering | throbbing sensation in brain and eye | 1 | 1 | 5.00 | 5 |
| cTBS (Fp1) | Eye/watering | watering eye (left eye) | 1 | 1 | 8.00 | 8 |
| cTBS (Fp1) | Other | Agitated | 1 | 1 | 10.00 | 10 |
| cTBS (Fp1) | Other | Tingling sensation | 1 | 1 | 0.00 | 0 |
| cTBS (Fp1) | Other | at the highest level, the poking sensation | 1 | 1 | 6.00 | 6 |
| cTBS (Fp1) | Other | stiffness, clenching self | 1 | 1 | 3.00 | 3 |
| cTBS (Fp1) | Other | tooth throbbing | 1 | 1 | 7.00 | 7 |
| cTBS (Fp1) | Other | tooth throbbing (at the highest amplitude) | 1 | 1 | 7.00 | 7 |
| cTBS (Fp1) | Pain/tapping sensation | butt a little sore | 1 | 1 | 2.00 | 2 |
| cTBS (Fp1) | Pain/tapping sensation | claustrophobia due to pillow around head and tapping of coil on forehead | 1 | 1 | 8.00 | 8 |
| cTBS (Fp1) | Pain/tapping sensation | cluster of shocking on scalp | 1 | 1 | 6.00 | 6 |
| cTBS (Fp1) | Pain/tapping sensation | highest setting pain, shocking kind of feeling | 1 | 1 | 5.00 | 5 |
| cTBS (Fp1) | Pain/tapping sensation | more intense tapping sensation, like poking | 1 | 1 | 7.00 | 7 |
| cTBS (Fp1) | Pain/tapping sensation | pricking feeling on scalp when amplitude increased | 1 | 1 | 3.00 | 3 |
| cTBS (Fp1) | Pain/tapping sensation | scalp/brain pain | 1 | 1 | 5.00 | 5 |
| cTBS (Fp1) | Teeth/jaw sensation | Teeth chattering | 1 | 1 | 2.00 | 2 |
| cTBS (Fp1) | Teeth/jaw sensation | Teeth tingling feeling | 1 | 1 | 2.00 | 2 |
| cTBS (Fp1) | Teeth/jaw sensation | pulsing sensation on 1 or 2 teeth back from upper left canine | 1 | 1 | 4.00 | 4 |
| cTBS (Fp1) | Teeth/jaw sensation | teeth vibration | 1 | 1 | 6.00 | 6 |
| cTBS (Fp1) | Teeth/jaw sensation | throbbing, pulsing sensation on 1 or 2 teeth back from upper left canine | 1 | 1 | 3.00 | 3 |
| cTBS (Fp1) | Vision change | alteration in vision (vision went black for a little bit) | 1 | 1 | 2.00 | 2 |

### Supplementary Figures

#### eFigure 1. Within-Day Session Trend in Maximum Tolerated Intensity

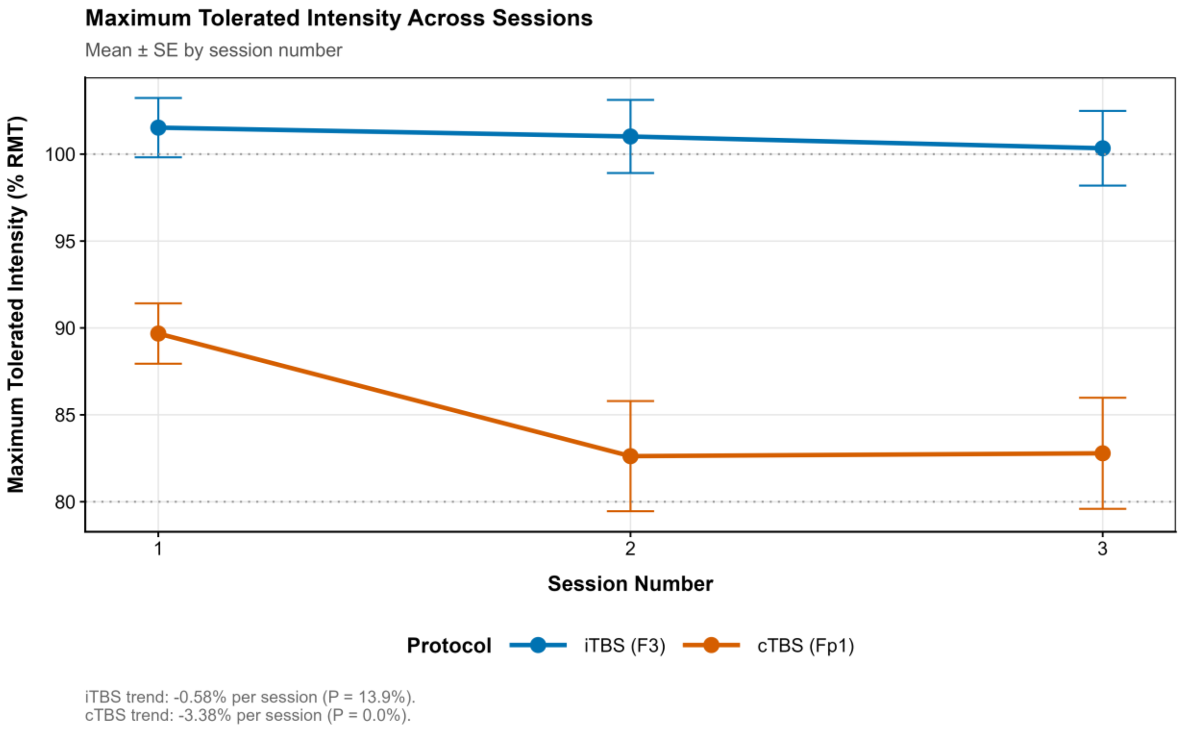


*Mean maximum tolerated intensity across the 3 within-day sessions for each protocol, with 95% credible intervals from the session-effect model.*

#### eFigure 2. Demographic Subgroup Forest Plot

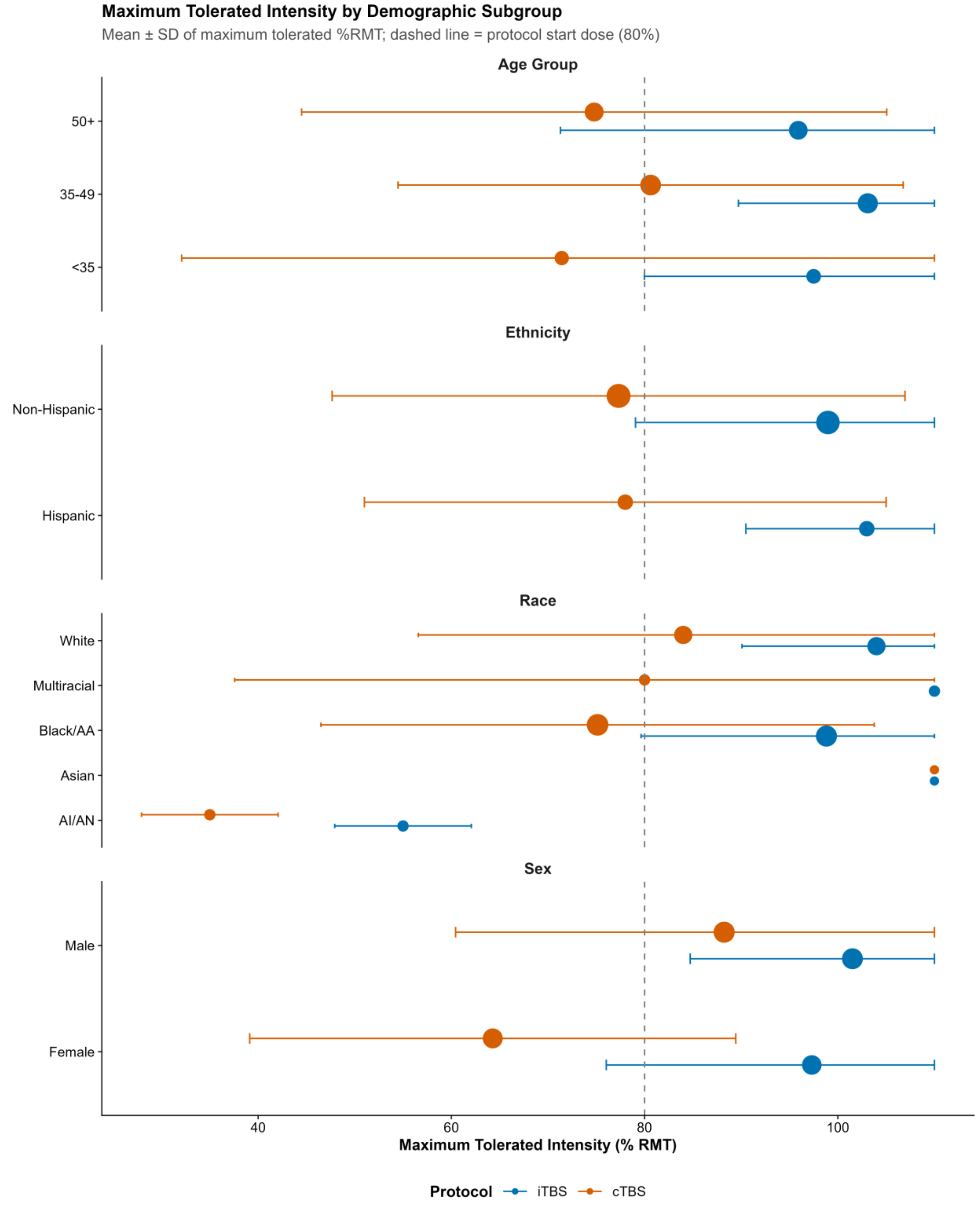


*Forest plot showing mean maximum tolerated intensity (+/- SD) by sex, age group, race, and ethnicity, stratified by protocol. Point size is proportional to subgroup sample size. Dashed line marks the protocol starting dose (80% RMT).*

#### eFigure 3. Posterior Predictive Check

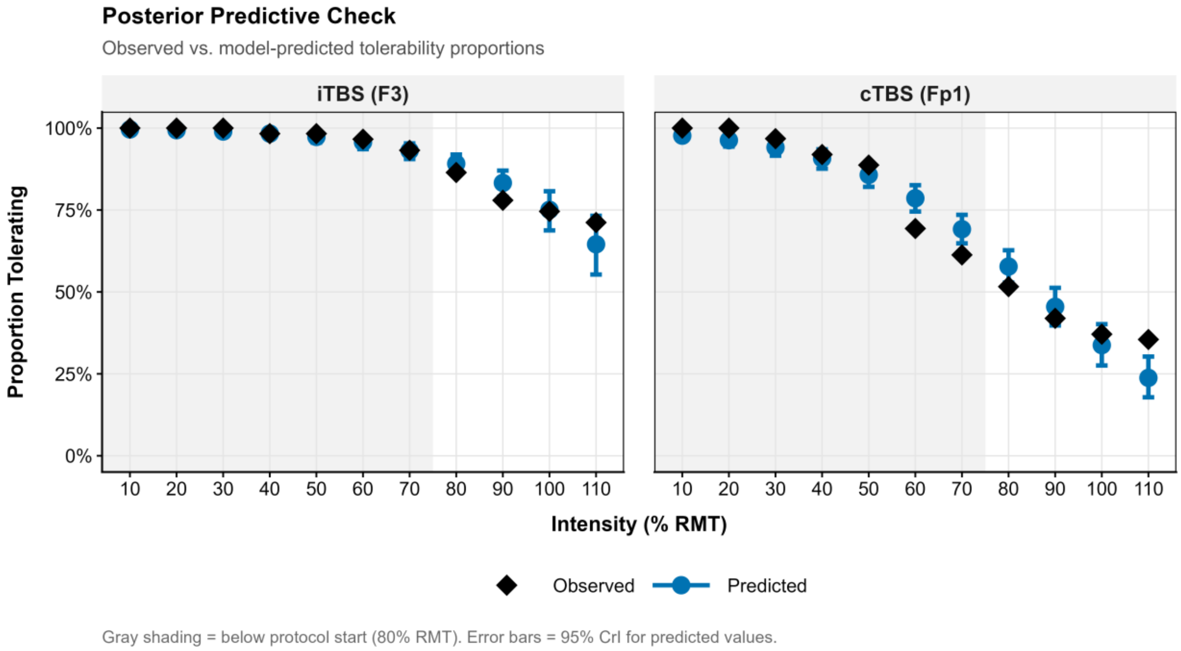


*Observed tolerability proportions overlaid on posterior model predictions at each intensity, verifying adequate model fit for both protocols.*

#### eFigure 4. Effect of Stimulation Order on Tolerability

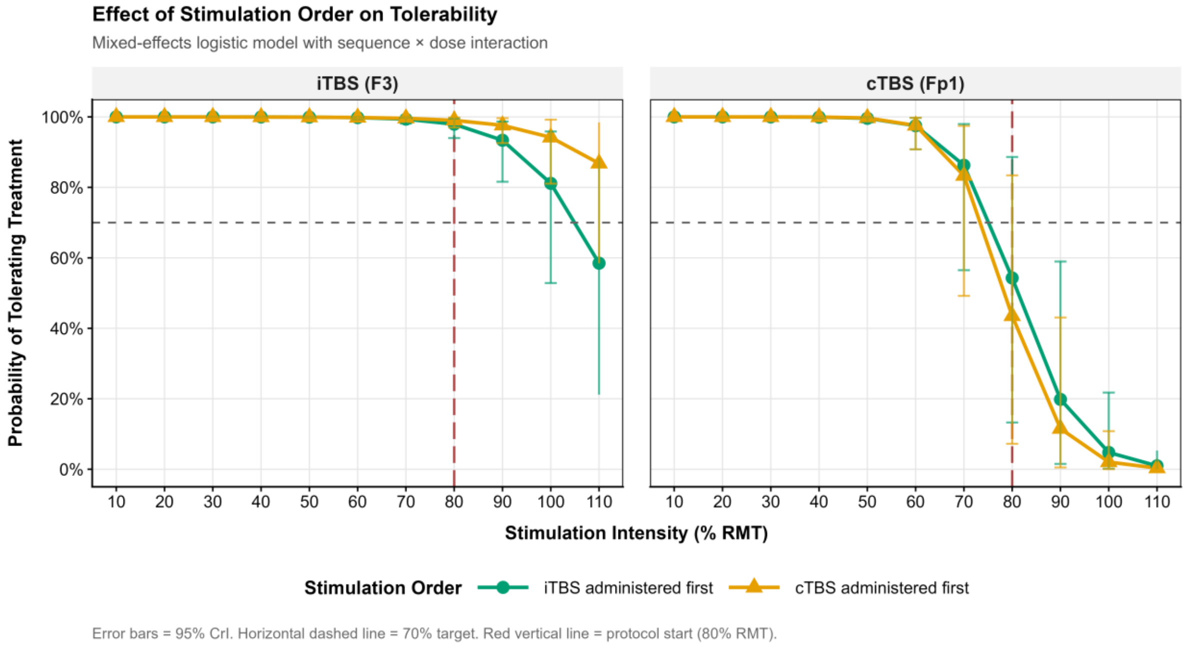


*Mixed-effects sequence-by-dose model results for iTBS and cTBS, showing dose-response curves stratified by treatment sequence (iTBS-first vs cTBS-first) with 95% credible intervals. No clinically meaningful sequence effects were observed for either protocol.*

**eFigure 5. Immediate Post-Session Resolution Among Symptoms Present During TMS**

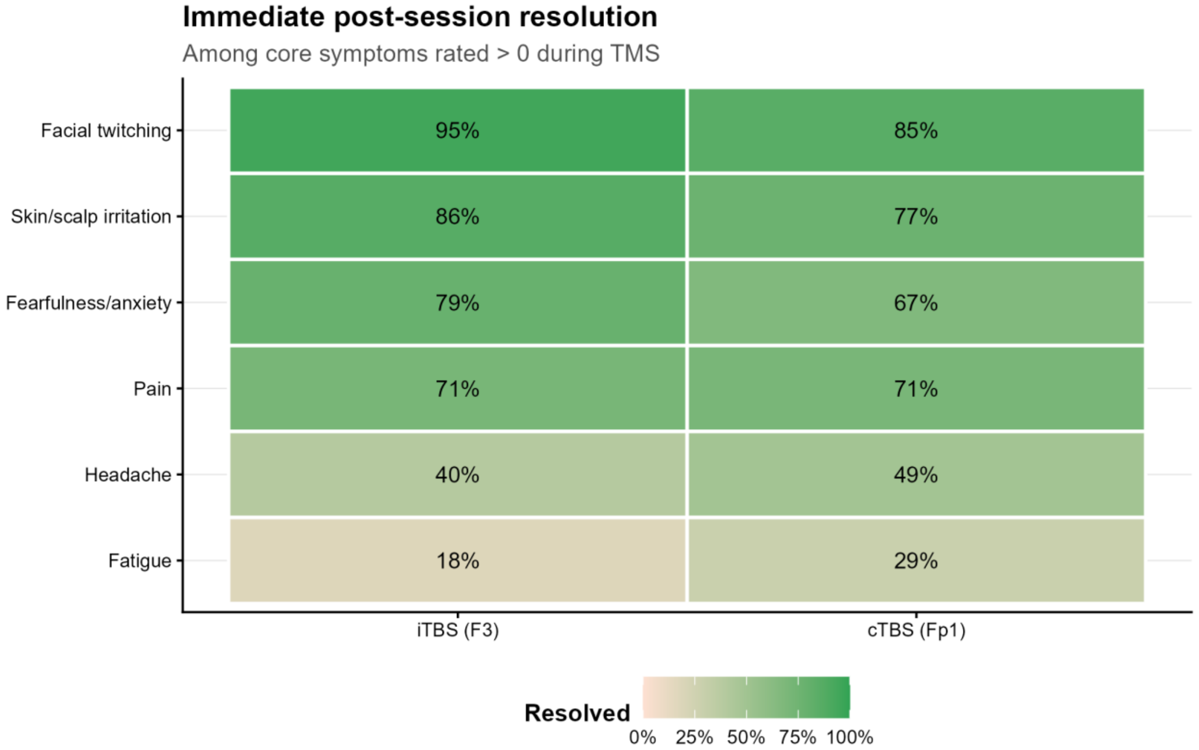


*Heatmap shows the percentage of symptom occurrences present during TMS (rating >0) that resolved by the immediate post-session assessment (rating = 0), stratified by protocol. Cells are shown for core symptoms with during-TMS occurrences in the corresponding protocol; darker green indicates higher resolution.*
